# Beyond the classical plasma secretome: genetic architecture and disease associations of the expanded human plasma proteome in 13,445 Europeans

**DOI:** 10.64898/2026.07.23.26358667

**Authors:** Solène Cadiou, Eva König, Alessia Mapelli, Giulia Pontali, Dariush Ghasemi-Semeskandeh, Michele Filosi, Brian R Ferolito, Michela C Massi, Gianmauro Cuccuru, Xiyun Jiang, Giovanna Winicki, Albert Navarro-Gallinad, Kai Gravel-Pucillo, Nicola Pirastu, Arianna Landini, Sodbo Sharapov, Johannes Rainer, Martin Gögele, Rebecca Lundin, Deborah Mascalzoni, Roberta Biasiotto, Hagen Blankenburg, Alessandro De Grandi, Clemens Egger, Lis Arend, Fabian Woller, Francesca Ieva, Nicole Soranzo, Kelly Cho, John Michael Gaziano, Luisa Zuccolo, Francisco S Domingues, Cristian Pattaro, John Danesh, Peter P Pramstaller, Alexandre Pereira, Emanuele Di Angelantonio, Christian Fuchsberger, Claudia Giambartolomei, Adam S Butterworth

## Abstract

Circulating plasma proteins are key biomarkers and therapeutic targets, now measurable at scale through high-throughput technologies, yet whether expanding proteomics platforms beyond the classical plasma secretome enhances genetic discovery and causal inference remains poorly understood. Here, we use an expanded SomaScan 7k platform to map the genetic architecture of a broader segment of the plasma proteome and to evaluate how proteome expansion affects pQTL discovery, causal inference and therapeutic target prioritisation. After quality control, we analysed 7,144 aptamers targeting 6,267 proteins in the harmonised dataset of two European cohorts: INTERVAL (n = 9,251 participants) and CHRIS (n = 4,194), and conducted genome-wide pQTL association analyses followed by meta-analysis. We identified 7,870 significant pQTLs (P-value < 1.26 × 10^−11^; 1,784 *cis*, 6,086 *trans*), of which 2,704 (34%) associations were not reported in five prior large-scale pQTL studies. Newly assessed proteins, which accounted for 53% (1,422/2,704) of the novel associations, were less likely to harbour *cis*-pQTLs associations (∼15%) than those in the previous platform version (∼28%), consistent with their lower expected plasma concentrations and predominantly intracellular localisation. Colocalization analyses revealed widespread sharing of genetic signals across proteins and characterised 22 pleiotropic *trans*-regulatory hotspots accounting for 68% of all *trans*-pQTLs. Through two-sample Mendelian randomization analyses on 2,003 phenotypes from the Million Veteran Program, UK Biobank, and FinnGen (combined N > 1.2 million), we identified 6,340 genetically supported protein–trait associations, highlighting disease mechanisms and potential therapeutic opportunities beyond currently drug-targeted circulating proteins. Together, these findings provide a systematic view of the genetic architecture of the expanded plasma proteome and demonstrate that plasma proteome expansion reveals genetically anchored disease biology beyond the classical secretome, while exposing inherent biological and technical constraints of studying low-abundance intracellular proteins in circulation.

## Introduction

Genome-wide association studies (GWAS) using high-throughput proteomic platforms such as Olink and SomaScan have provided key insights into the genetic architecture of plasma levels of up to 5,000 proteins, revealing novel mechanistic pathways for complex traits and diseases.^1–4^ These studies have shown that plasma protein levels are substantially heritable, shaped by both *cis*- and *trans*- acting genetic variants, and have established protein quantitative trait loci (pQTLs) as a scalable framework for drug target validation. In particular, Mendelian randomization using pQTL studies provide a scalable framework to prioritize therapeutic targets and identify repurposing opportunities across diseases.^5,6^

As assay platforms continue to expand proteome coverage, it remains unclear whether assay performance, detection power, and genetic architecture are maintained for newly added proteins. For example, initial analyses of the Olink Explore HT platform, which expands from 3,000 to 5,000 target proteins, suggest that the newly assayed proteins are measured less reliably.^7^ Whether this expanded coverage meaningfully extends causal inference to previously inaccessible biological pathways, and at what cost to statistical power and interpretability, has not been systematically evaluated.

Here, we report a large-scale pQTL meta-analysis of 7,144 proteins measured using the expanded SomaScan 7k platform in 13,445 participants from two European cohorts, systematically comparing newly measured proteins with those on the previous 5k protein version. This design allows to evaluate how proteome expansion affects pQTL discovery, causal inference, and therapeutic target prioritisation, while extending the understanding of the genetic architecture of plasma proteome in Europeans.

## Results

### Genetic architecture of the expanded human plasma proteome

Genome-wide association analyses of plasma protein levels measured using the SomaScan 7k platform in 13,445 European-ancestry participants from the INTERVAL^8^ (n = 9,251) and CHRIS^9,10^ (n = 4,194) studies revealed extensive genetic regulation across the expanded proteome. After quality control and filtering of poorly detectable aptamers, the associations between 8,613,862 variants (MAC>10 in both studies) and 7,144 aptamers targeting 6,267 proteins were tested with cohort-specific covariate adjustment (**Methods, Supplementary Table 1, SM4**). Genomic inflation was well controlled across all analyses (median λGC = 1.005 (range: 0.975–1.094)).

We identified 7,870 significant protein quantitative trait loci (pQTLs regional associations; aptamer–locus pairs; see **Methods, SM6**) after multiple testing correction (P-value < 1.26 × 10^−11^), comprising 1,784 (22.7%) *cis* - signals located within +-500 kb of the Transcription Start Site of the encoding protein - and 6,086 (77.3%) *trans* associations (**Figure 1a**; **Supplementary Table 2**). Overall, 4,361 proteins (69%) had at least one pQTL, and 1,781 proteins (28.4%) had at least one *cis*-pQTL. *Cis* associations were broadly distributed across the genome, whereas *trans* associations clustered in discrete pleiotropic regulatory hotspots (**Figure 1a,b**).

**Figure 1.**
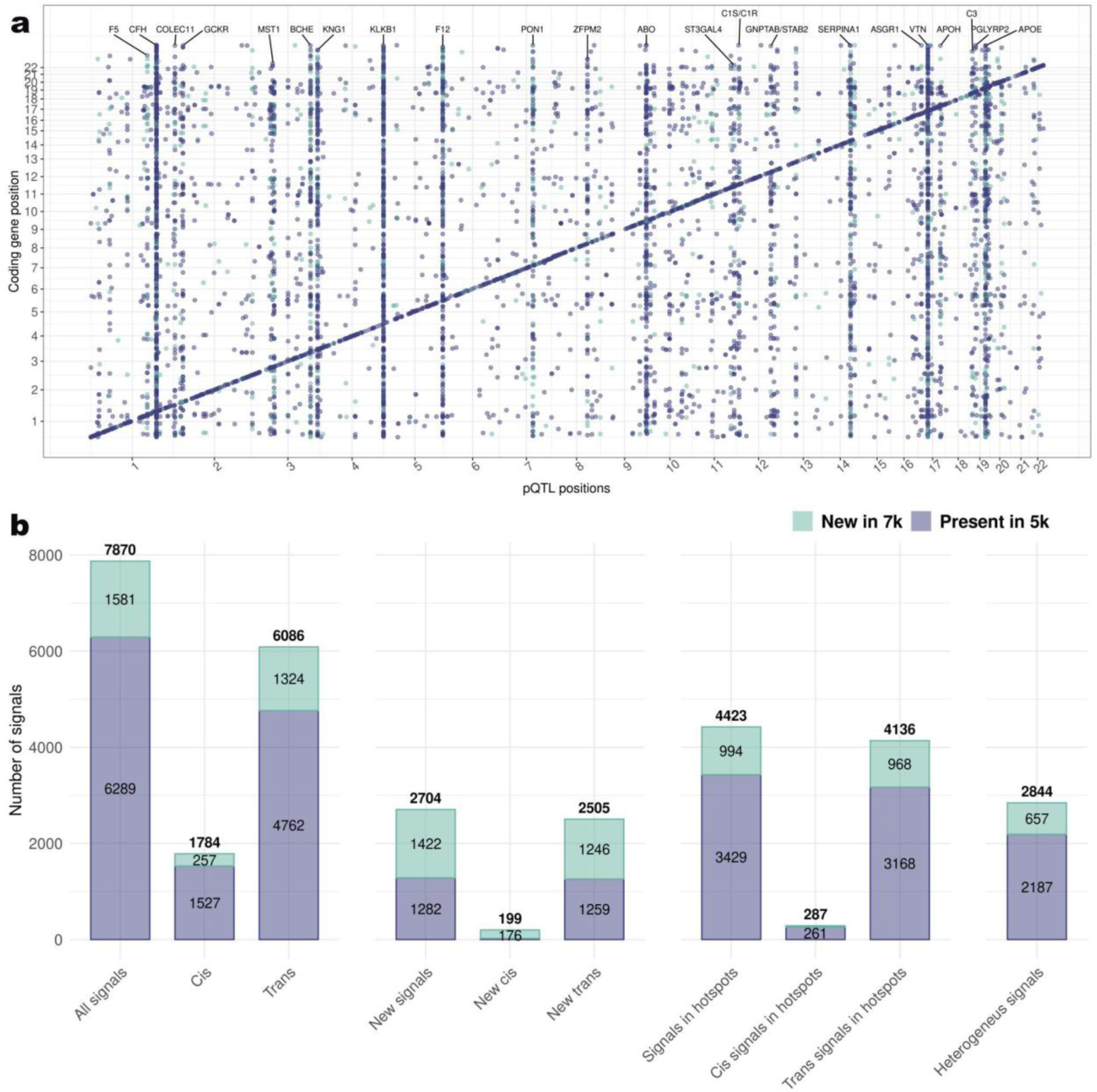
Overview of the genetic architecture of the plasma proteome in the SomaScan 7k meta-analysis. Characteristics of significant regional associations from the meta-analysis of 7,144 aptamers, categorized according to whether the corresponding protein was newly assessed in the SomaScan 7k assay. (**a**) Genomic positions of all 7,870 significant regional associations and the corresponding coding genes, colored by whether the protein was newly assessed or not in the 7k assay. For each hotspot (highly pleiotropic regions), one gene name was chosen. (**b**) The bars show the proportion of newly vs previously assayed aptamers across subsets of regional associations (loci-aptamer level).

Comparison with five large European-ancestry pQTL studies^1–4,11^ (**Methods**; **SM13**) showed that 2,704 signals (34.3%) were not reported previously, including 199 *cis* and 2,505 *trans* associations. Most of these novel signals (1,972; 72.9%) were located within *trans*-regulatory hotspot regions (**Figure 1b**). As expected, most novel *cis* associations (176 of 199; 89%) involved one of the 1,682 proteins not included on the previous version of the SomaScan platform.

To characterize locus-level complexity, we performed conditional association analysis, identifying 12,095 conditionally independent variants across the 7,870 significant regional associations (**Supplementary Table 3**; **Supplementary Figure 1**). These included 2,983 additional independent signals at *cis* loci (4,767 total) and 1,243 (7,329 total) at *trans* loci, indicating widespread allelic heterogeneity beyond index variants.

Among the 1,784 *cis*-pQTLs, 700 (39%) had at least one conditionally independent association where the index variant, or a strong proxy (r² > 0.8), was a protein-altering variant (PAV) within the gene encoding the target protein (**Methods**; **SM20**; **Supplementary Table 3**). At 244 *cis*-pQTLs (14%), all conditionally independent signals included a PAV. Because PAVs can alter aptamer binding affinity independently of protein abundance, these loci require careful interpretation, as apparent protein differences may reflect sequence-dependent assay effects in addition to, or instead of, true biological variation in circulating protein levels. Accordingly, we considered these signals as potentially epitope-sensitive in downstream causal inference analyses and systematically annotated them to distinguish likely abundance-driven from potentially binding-driven associations.

Proteins newly introduced in the SomaScan 7k platform had less *cis*-pQTLs than proteins already present in the earlier 5k SomaScan version (15% versus 28%), whereas differences in the probability of detecting any pQTL (*cis* or *trans*) were more modest (57.8% versus 63.6%; **Figure 1b; Supplementary Figure 2**). These observations motivated further analyses to determine whether reduced *cis*-pQTL discovery among newly measured proteins predominantly reflects biological properties of the expanded proteome or technical characteristics of the assay.

### Reduced *cis*-pQTL discovery among expanded proteome reflects the introduction of intracellular, low-abundance proteins

The expansion from the 5k to the 7k SomaScan platform substantially altered the biological composition of the measured proteome. Of the 6,381 proteins targeted by the SomaScan 7k platform, 1,682 (26.4%) were newly introduced relative to the earlier 5k version (**Supplementary Table 1; Supplementary Figure 3**). These newly added proteins were strongly enriched for intracellular localization and depleted for classical secreted proteins (**Supplementary Figure 3a**). Consistent with this shift, newly measured proteins were predicted from Human Protein Atlas annotations to occur at markedly lower circulating concentrations than proteins already present on the 5k platform. This pattern suggests that the 7k expansion extended coverage beyond the traditional plasma secretome towards lower abundant proteins likely entering circulation through cellular turnover, vesicular release or tissue leakage (**Supplementary Figure 3g**).

Despite these biological differences, assay precision metrics and limits of detection were comparable between newly added and previously assayed proteins across cohorts and batches (**Supplementary Figures 4 and 5, Supplementary Figure 3c–e, SM2**). This indicates that even if the platform expansion changed the biological spectrum of proteins measured rather and thus what can be observed, the underlying analytical performance of the assay was maintained.

To quantify determinants of *cis*-pQTL discovery, we fitted logistic regression models incorporating protein biological annotations and assay performance metrics (**Supplementary Table 4, SM10**). Protein biological characteristics were the strongest predictors of *cis*-pQTL detection. Secreted proteins were more than four times more likely to harbour a *cis*-pQTL than intracellular proteins (OR = 4.25, P-value = 3.69 × 10^−106^), and higher expected circulating concentration independently increased *cis* discovery (OR per log_−10_ unit = 1.53, P-value = 6.58 × 10^−65^). In contrast, assay precision (coefficient of variation) and detectability effects on *cis*-pQTL discovery were modest (descriptive stratification of proteins by *cis*-pQTL status is shown in **Supplementary Figure 2**).

Interestingly, allelic heterogeneity at *cis*-pQTL loci was also strongly linked to protein localization, with proteins exposed at or beyond the cell surface (i.e. membrane-bound and secreted) exhibiting markedly greater allelic complexity than intracellular proteins.

Although newly introduced proteins showed reduced *cis*-pQTL discovery in unadjusted analyses (**Figure 1**, OR = 0.71, P-value = 3.01 × 10^−8^), this difference was largely attenuated after accounting for protein localization, circulating concentration, and detectability (OR_adjusted_ = 0.3, P-value = 0.004), indicating that reduced *cis*-pQTL discovery among newly measured proteins is largely explained by their biological characteristics, particularly intracellular localization and lower expected circulating abundance, rather than reduced assay sensitivity. Indeed, because many of these proteins are thus expected to be present near the limits of plasma detectability, reduced inter-individual variation and limited statistical power may contribute to lower *cis*-pQTL detection, highlighting the intrinsic challenges of studying intracellular proteins in plasma rather than in their tissue of origin.

### *Trans*-pQTLs are dominated by pleiotropic regulatory hotspots, whereas *cis* architectures vary by protein localization

*Trans*-pQTLs were highly concentrated within a limited number of pleiotropic regulatory hotspots, defined as merged contiguous 500 kb windows enriched for overlapping signals (**Methods, SM11**). We identified 22 genomic regions that collectively accounted for 4,136 (68%) of all *trans* regional associations (**Figure 1a; Supplementary Table 5**).

Between-study heterogeneity was enriched within these *trans*-regulatory hotspots. Overall, 36% (2,844) of pQTL signals showed high heterogeneity across cohorts (I² >= 90), and 79% (2,236) of these highly heterogeneous signals were located within hotspot regions. Heterogeneity varied across hotspots, ranging from 95% of signals showing high heterogeneity to only 1% (**Supplementary Figure 6a**). Interestingly, a strong correlation was observed between proportion of heterogeneous signals and proportion of previously reported signals: the most heterogeneous hotspots were also the ones less confirmed in the literature, suggesting study-specific effects (**Supplementary Figure 6b)**. A manual curation of hotspot regions suggested several potential mechanisms underlying such widespread *trans* effects. A first class comprised loci containing genes mapping to coagulation pathways, including *APOH, F5, F12, KNG1/KLKB1, SERPINA1* and *VTN*. The hotspots involving these loci had the highest between-study heterogeneity and lowest proportions of replication in previous studies (**Supplementary Figure 6b)**, consistent with sensitivity to coagulation-related pre-analytical variation,^4,12^ although *in vivo* biological effects cannot be excluded. A second class included more reproducible hotspots containing genes mapping to lipid metabolism and hepatic clearance pathways, including *GCKR, APOE, BCHE, GNPTAB/STAB2* and *ASGR1*. Additional hotspots, including *ABO, ST3GAL4, COLEC11* (modification of cell surface properties) *C1R/C1S, C3*, *CFH*, (complement activation proteins), *MST1* and *PGLYRP2* (scaffold and carrier proteins) were consistent with immune, complement, or structural effects, whereas *ZFPM2* represented a transcriptional regulator locus potentially influencing broader blood-cell biology. Together, these observations suggest that *trans*-pQTL hotspots can arise through multiple mechanisms, including broad regulation of important biological pathways, plasma compositional effects, and sensitivity to sample handling.

*Cis*-regulatory architectures were more homogeneous, widely distributed across the genome and varied systematically with protein biology. Among aptamers with at least one *cis*-pQTL, newly introduced SomaScan 7k aptamers showed reduced allelic heterogeneity at *cis*-pQTL loci: 43.5% (111/255) harboured two or more conditionally independent *cis* variants compared with 60.6% (924/1,526) of aptamers present in the earlier platform (OR = 0.50, P-value = 4.7 × 10⁻⁷) (**Supplementary Table 4, SM16**). This difference was eliminated after adjustment for protein localization and expected circulating concentration (OR = 0.77, P-value = 0.07), indicating that simpler observed *cis* architectures among newly measured proteins may reflect reduced statistical power due to their lower concentration in plasma.

### Colocalization reveals shared and network-level genetic regulation of circulating proteins

Extensive sharing of genetic signals was observed across proteins. Genome-wide pairwise colocalization between conditionally independent pQTL signals whose 99% credible sets shared at least one variant identified 582,245 pairs with strong evidence of a shared causal variant (PP.H4 > 0.8), indicating widespread sharing of association signals (**Methods; SM14**).

Nearly all colocalizing pairs (99%, n = 576,307) involved two *trans*-pQTLs, consistent with extensive pleiotropic regulation at *trans* hotspots (99%, n = 569,048 where at least one of the *trans-*pQTLs belonged to a hotspot). Notably, 682 (38%) of 1,784 *cis*-pQTLs also colocalized with at least one *trans*-pQTL, providing the opportunity to identify likely effector genes at *trans*-regulatory loci. For example, a previously reported *trans*-pQTL for tenascin-C (1:960941:C:G; P-value = 8.3 × 10^-17^; seq.2728.62), a widely expressed extracellular matrix protein that plays a role in peripheral nervous system axon regeneration, colocalized strongly (PP.H4 = 0.99) with a novel *cis*-pQTL for a previously unmeasured protein, agrin (1:968226:A:C; P-value = 1.7 × 10^-131^; seq.15483.377), suggesting *AGRN* as a candidate effector gene at this locus (**Supplementary Figure 7**). Agrin is a heparan sulfate proteoglycan that is believed to be important in maintenance of the blood-brain barrier. Previous research has suggested that agrin loss strongly correlates with tenascin expression, supporting our genetic evidence that agrin may regulate tenascin.^13^

We identified a dense cluster of 13 robust colocalizing signals at the *MGAT5* locus indexed by 2:134966562:A:C (rs62165726). Of these, 11 are *trans*-pQTLs, either previously reported according to our literature review (signals for proteins encoded by *CSF1R, CR1, VCAM1, SIGLEC1 and LILRB1*) or novel (signals for proteins encoded by *MERTK, LRRC4 and LILRA1*). These 11 *trans*-pQTLs colocalize with 2 newly reported *cis* signals with aptamers targeting alpha-1,6-mannosylglycoprotein 6-beta-N-acetylglucosaminyltransferase A (*MGAT5*), a protein newly assessed in the 7k assay (seq.21813.171 and seq.21768.9, P-value = 1.31 x10^-20^ and 1.56 x 10^-20^ respectively). *MGAT*5 belongs to the glycosyltransferase family and is involved in the regulation of the biosynthesis of glycoprotein oligosaccharides.^14^ As alterations of oligosaccharides on cell surface glycoproteins can cause significant changes in adhesion properties and in migratory behavior of cells, our genetic evidence nominates *MGAT5*-mediated glycosylation as a plausible mechanism underlying this *trans*-pQTL cluster. In support of this interpretation, experimental studies have shown *MGAT5*-dependent N-glycosylation can affect eosinophil recruitment and adhesion-related phenotypes involving VCAM1^15^ while glycan-binding studies show that complex sulfated N-glycans contribute to ligand recognition across the human Siglec family, including SIGLEC1.^16^ However, direct MGAT5-dependent regulation of these *trans*-pQTL proteins, including SIGLEC1, remains to be established.

Beyond linking *trans* signals to candidate effector genes, colocalization analyses also revealed frequent multi-protein regulation at *cis* loci. We identified 935 pairs of colocalizing *cis*-pQTLs of which 105 target different proteins, involving 154 proteins. These often-affected members of the same gene family, including *C1R/C1S*, *GSTM1/GSTM3/GSTM4* and *MMP1/MMP13*, demonstrating that *cis*-pQTL loci frequently harbour multi-target regulatory effects, complicating straightforward attribution of causality to a single protein and underscoring the need for caution when interpreting regional signals in downstream analyses. As proteomic breadth increases, such multi-target regulatory structure becomes more visible, underscoring that *cis*-pQTLs should not be assumed to uniquely tag one protein even at apparently simple loci.

This shared regulation was particularly evident within dense *trans*-regulatory hotspots. To determine whether the 22 hotspot loci reflected single highly pleiotropic variants or clusters of multiple independent pleiotropic effects, we built networks of robust (LD > 0.8 between independent index SNP) colocalization signals (**Methods; SM17** and **Supplementary Table 5**). At some loci, *trans* regulation was consistent with a single dominant driver. For example, at the *GCKR* locus on chromosome 2, 75 *trans*-pQTL signals aligned with the well-characterized index SNP p.Pro446Leu (rs1260326; 2:27730940:T:C), indicating a single pleiotropic mechanism underlying coordinated regulation across multiple proteins (**Figure 2a, Supplementary Figure 8a**).

**Figure 2.**
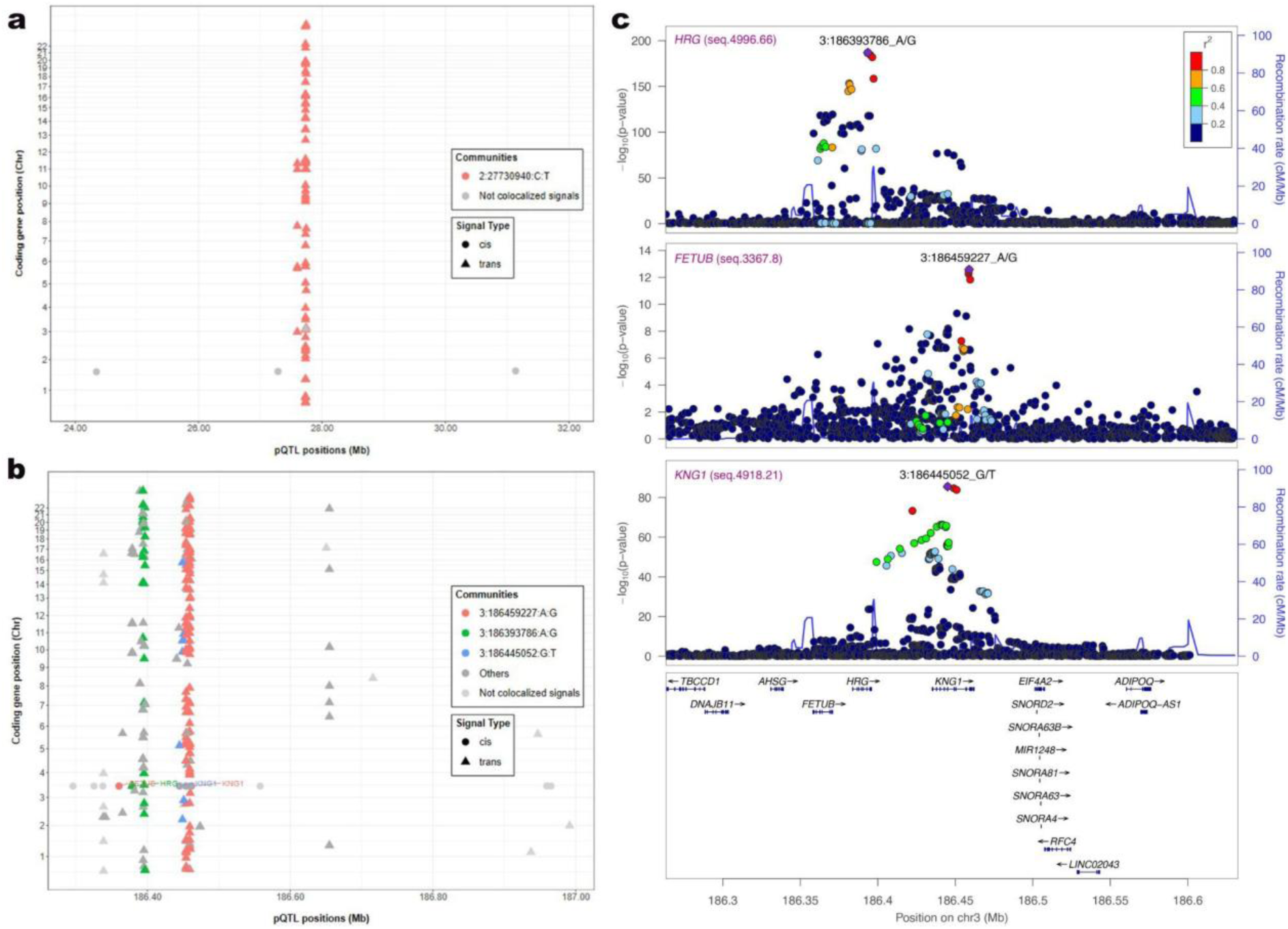
Regulatory hotspots shape *trans* architecture of the plasma proteome. (**a**) Example of a homogeneous hotspot on chromosome 2 (*GCKR* locus), where 75 *trans*-pQTL signals colocalize with the well-characterized index variant rs1260326 (2:27730940:C:T). The fully connected colocalization network (**Supplementary Figure 13**) indicates a single pleiotropic mechanism driving coordinated changes across multiple proteins. (**b**) Example of a heterogeneous hotspot on chromosome 3, where multiple distinct clusters of *trans*-pQTLs are linked to different *cis*-regulatory variants within the *FETUB/HRG/KNG1* region. The plot illustrates three main colocalization communities (see network in **Supplementary Figure 13b**). (**c**) Regional association plot of a heterogeneous hotspot on chromosome 3, with examples of three distinct *cis*-pQTLs that each colocalize with many *trans-*pQTLs. Top, LocusZoom plot of *HRG* aptamer (seq.4996.66). Middle, LocusZoom plot of *FETUB* aptamer (seq.3367.8). Bottom, LocusZoom plot of *KNG1* aptamer (seq.4918.21). Data used to plot the associations of the variants with each aptamer is based on region-specific conditional analysis to isolate the relevant association signal with the plots bounded to a fixed region (*chr3:186,263,141-186,632,505* (GRCh37)) for the visualisation purposes. The pairwise LD between variants was computed based on the genotype data of the participants with proteomic data in the INTERVAL study. The plot was generated with LocusZoom v1.4.

In contrast, other hotspots exhibited more complex architectures. A gene-dense region on chromosome 3 (186-187Mb) contained 327 conditionally independent pQTLs, including *cis-*pQTLs for 7 proteins, with some having multiple independent signals (e.g. HRG (9 signals), FETUB (8), AHSG (5), and ADIPOQ (4)). Many of the *trans-*pQTLs clustered into three distinct communities: (a) a cluster of more 115 *trans*-pQTLs with a *cis*-pQTL for FETUB and 2 *cis*-pQTL for KNG1 (3:186459227:A:G); (b) a cluster of 10 *trans*-pQTLs with another *cis*-pQTL for KNG1 (3:186445052:G:T); (c) a cluster of more than 30 *trans*-pQTLs with a *cis*-pQTL for HRG (3:186393786:A:G) (**Figure 2b,c; Supplementary Figure 8b**). These partially distinct colocalization patterns centred on different *cis*-pQTL signals suggest that the hotspot is unlikely to be explained by a single shared LD structure alone and instead reflects multiple overlapping *trans*-regulatory components. Together, these examples illustrate that a small number of genomic regions dominate *trans*-protein regulation, but that these hotspot regions vary from single master regulator variants (as seen for *GCKR*) to more complex regional architectures involving multiple partially independent *cis* and *trans* association patterns, as seen for chromosome 3.

### pQTLs show strong transcriptional concordance with *cis*-eQTLs but also reveal multi-gene regulatory complexity and tissue-specific discordance

We used *cis*-eQTLs from GTEx^17^ v8 (European ancestry) to assess concordance between observed plasma protein genetic regulation and gene expression across 49 tissues.

We first assessed whether conditionally independent *cis*-pQTL signals were associated with gene expression of the protein-coding gene using an eQTL lookup approach. Among the 4,767 independent *cis*-pQTL index variants, 3,234 (68%) were also a significant *cis*-eQTL (FDR ≤ 5%) for the corresponding protein-encoding gene in at least one tissue. Effect-direction concordance for the same gene was also generally strong, with 27,065 (78%) of 34,528 eQTL-pQTL pairs showing directional consistency across tissues. We then performed formal colocalization between conditionally independent pQTL signals with *cis*-eQTLs (**Methods; SM18** and **Supplementary Table 3**). Overall, 6,876 (87%) of 7,870 pQTLs (87%) colocalized with at least one gene in at least one tissue. Focusing on *cis*-pQTL regions, the gene encoding the protein colocalized in at least one tissue for 1,214 (68%) of 1,784 regions.

We observed some consistent patterns across tissues, suggesting that plasma protein levels reflect gene expression regulation predominantly in specific tissues, thereby underscoring the importance of multi-tissue analysis. Indeed, consistently with previous reports, effect direction concordance between *cis*-pQTLs and *cis*-eQTLs for the same gene was generally high (0.67-0.86, median=0.79) but varied substantially across tissues, with the highest concordance observed in small intestine (0.86) and liver (0.85), which directly contribute to protein secretion in blood, and lower concordance in brain tissues (0.67-0.82, median=0.75), as the blood-brain barrier restricts the passage of proteins (**Supplementary Figure 9a**). Systematic patterns of concordance or discordance were also observed directly at the level of index variants: 1,707 of 3,234 *cis*-pQTLs (53%) showed consistent effect direction with a significant *cis*-eQTL in all tissues, 536 *cis*-pQTLs (17%) showed opposing effect direction in all tissues, while the remaining 991 *cis*-pQTLs (31%) showed a mix of consistent and opposing effect direction across tissues. An example of systematic discordance between transcript and protein levels is the *cis*-pQTL 11:72497462:C:T with the newly assessed aptamer seq.23569.53 (encoding *STARD10*, P-value = 1.2 × 10^-57^), which had directionally discordant *cis-*eQTLs in all tissues with a significant eQTL, suggesting inverse regulation of plasma protein levels and intracellular transcript levels (**Supplementary Figure 10a, Supplementary Table 3**). Furthermore, tissue-specific variability in effect direction is exemplified by the *cis*-pQTL 2:112858122:C:T with the newly assessed aptamer seq.21891.31 (encoding *FBLN7*, P-value = 5.1 × 10^-35^), which displayed concordant effects in artery and adipose tissues, thyroid and skin, but discordant effects in brain tissues (**Supplementary Figure 10b, Supplementary Table 3**), highlighting tissue-specific regulatory complexity.

For *cis*-eQTL colocalization, we also observed a biologically plausible pattern of tissue-specific enrichment: colocalization of the protein-coding gene with its *cis*-pQTLs occurred most frequently in whole blood (1.7x enriched), adipose, and muscle tissues, all tissues relevant to protein secretion into blood, while brain, possibly due to the blood-brain barrier, and kidney cortex (4.1x depleted) showed fewer colocalizations (**Supplementary Figure 9f**, **SM 18.2**). While this analysis did not consider the effect of different sample sizes for the GTEx tissues, it nevertheless suggests that tissues with less colocalization enrichment may benefit from tissue-specific proteomic studies to enhance our understanding of genetic regulation of proteins.

While lookup, direction concordance and colocalization have shown a strong overall concordance between *cis*-pQTL and *cis*-eQTL, eQTL colocalization was not observed for 32% of *cis*-pQTL loci. This may reflect regulatory mechanisms not mediated through steady-state transcript abundance, including post-transcriptional or secretion-related effects, but may also partly arise from limited statistical power or incomplete tissue representation in currently available eQTL datasets. Among the 244 *cis* regions where all conditionally independent signals involved PAVs, 84 (34%) showed no colocalization with the encoding gene in any GTEx tissue, similar to the proportion observed across all *cis*-pQTL loci. Thus, absence of colocalization is not enriched among PAV-driven signals and cannot be attributed solely to sequence-dependent assay effects. Rather, both PAV and non-PAV *cis*-pQTLs appear to arise from heterogeneous mechanisms, including coding, post-transcriptional, or transcriptional effects that may not be fully captured by steady-state eQTL data.

We also observed that colocalization was frequently non-specific to the gene encoding the protein: 89% of the colocalizing *cis*-pQTLs also showed colocalization with at least one other gene of the *cis* region in the same or a different tissue. No difference was observed when we stratified the analysis on newly vs previously assayed proteins. Interestingly, all the 415 *cis*-pQTLs that did not colocalize with the protein-encoding gene showed colocalization with at least one other gene (**Supplementary Figure 9c**). This can represent at least partially a statistical artefact, as the probability of colocalization increased with the size of the region and the number of genes tested (**Supplementary Figure 9d-e**) (additionally, for 155 *cis*-pQTLs (9%), the gene encoding the protein was not available in the GTEx data). However, it also underscores that, as demonstrated in our *cis*-pQTL/*cis*-pQTL colocalization analyses, *cis*-pQTL loci can harbour multi-target regulatory effects, complicating the interpretation of downstream analyses.

Finally, *cis*-eQTL colocalization analyses also highlight complex signal-sharing patterns with *trans*-pQTLs, where we observed widespread colocalization with multiple genes across loci and tissues. We found that only 1852 (30%) *trans*-pQTL regions had exactly one colocalizing gene, while 3395 (56%) had multiple colocalizing genes within at least one tissue (**Supplementary Figure 9g**). The median number of colocalizing genes per *trans*-pQTL loci was still lower than *for cis*-pQTL loci (respectively 2 (IQR: 1–5) and 4 (IQR: 2–7)).

Together, these findings indicate that while many *cis*-pQTLs align with transcriptional regulation of the encoding gene —particularly in secretion-relevant tissues—this concordance is not universal, and both *cis*- and *trans*-pQTL loci frequently exhibit heterogeneous and multi-gene regulatory architectures. Plasma protein regulation therefore reflects diverse mechanisms beyond steady-state transcript abundance, highlighting the layered nature of proteomic genetic effects.

### Proteome expansion broadens the landscape of genetically supported disease biology

To assess the translational relevance of these findings, we next performed systematic Mendelian randomization (MR) analyses to identify genetically supported links between circulating proteins and disease-relevant traits.

Starting from the 1,548 proteins showing at least one significant *cis* associations, we performed two-sample MR using 4,712 conditionally independent *cis-*pQTL instruments across 1,775 aptamers (corresponding to 1,543 proteins) against 2,003 phenotypes including disease diagnoses, clinical biomarkers and medication use from large-scale GWAS meta-analyses of the Million Veteran Program (MVP)^18^, UK Biobank (UKBB),^19^ and FinnGen^20^ biobanks including more than 1.2 million European-ancestry participants^6^ (**Supplementary Table 6**). MR exposures were defined as unique aptamer/protein-encoding gene pairs for which relevant *cis*-pQTL instruments were available (**Methods, SM19**).

At a Bonferroni-corrected threshold (P-value < 1.42 x 10^-8^, i.e. 0.05 / 3,497,303), we identified 7,591 significant triplets (aptamer/protein-encoding gene/phenotype), implicating 574 unique proteins (corresponding to 574 genes), 643 phenotypes, and 6,340 unique gene–phenotype pairs (**Figure 3a**, **Supplementary Table 7**). The higher number of triplets compared with unique gene–phenotype pairs reflects redundancy at both mapping steps: multiple aptamers may target the same protein, and multiple proteins may map to the same gene; consequently, distinct aptamer–protein–phenotype triplets can correspond to the same underlying gene–phenotype association. Among the unique gene-phenotype pairs, 1,768 were newly identified in the present study compared to previous work testing the same list of phenotypes with previously assessed eQTL and pQTL.^6^

**Figure 3.**
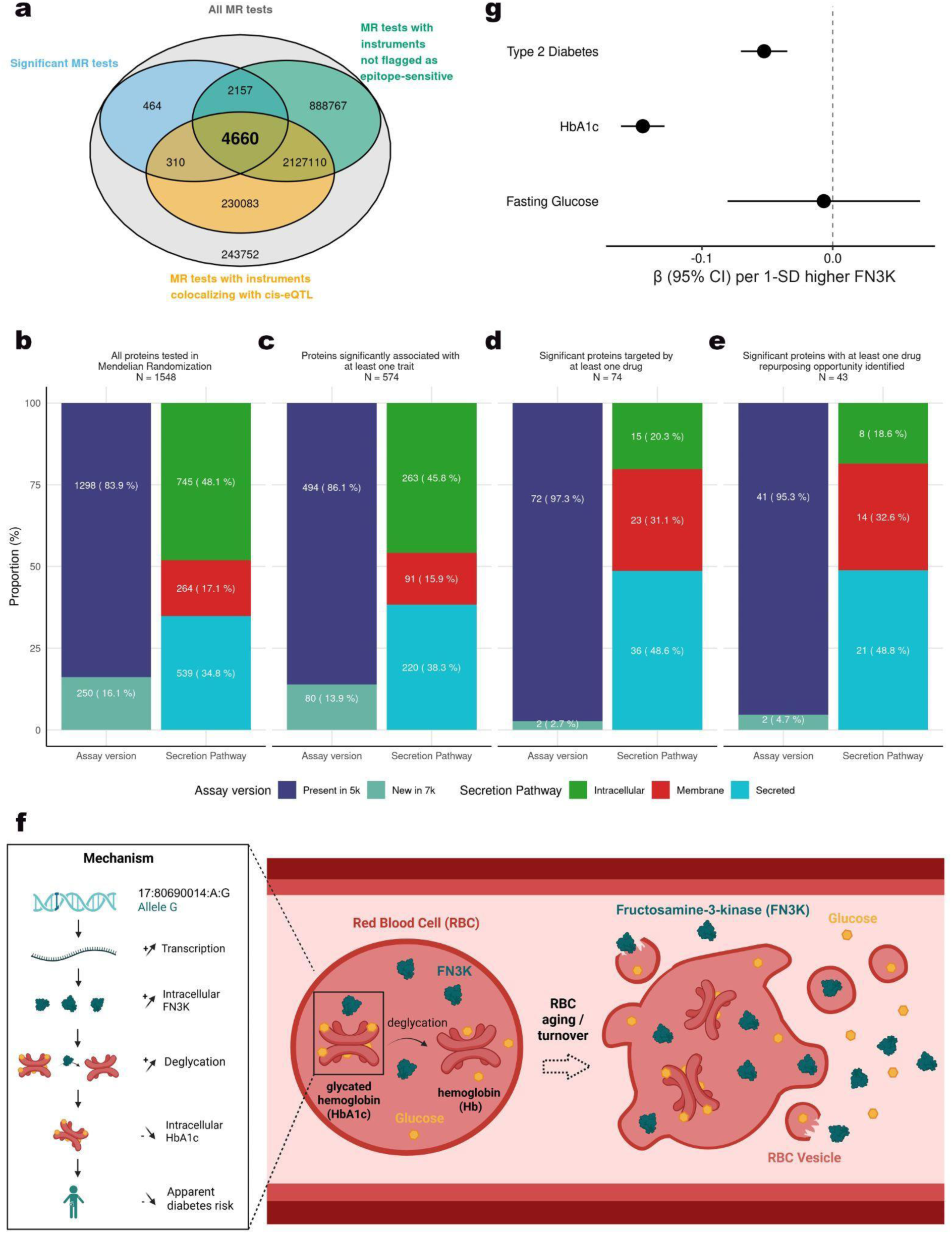
Genetically supported circulating protein targets across the therapeutic development spectrum. Systematic two-sample Mendelian randomization (MR) was performed for 1,548 proteins (corresponding to 1,784 aptamer-gene pairs) proteins and 2,003 phenotypes. After instrument matching, MR tests were effectively computed for 1,543 proteins (corresponding to 1,778 aptamer-gene pairs). (**a**) Venn Diagram characterizing all Mendelian Randomization tests performed (association of exposures defined as aptamer/protein-encoding gene with phenotypes, in grey) according to their significance after Bonferroni correction (blue), if the corresponding pQTL instrument was not flagged as possibly driven by epitope effect (green) and if it was colocalizing with the protein-encoding gene eQTL (orange). (**b-e**) Panels illustrate the progression from all proteins tested to proteins with increasing levels of therapeutic relevance. Proteins are stratified by secretion annotation (secreted, membrane, intracellular, other/missing) and whether uniquely measurable on the SomaScan 7k platform. Across panels, secreted proteins become progressively enriched from the fully tested proteome to drug-targeted and repurposing subsets, underscoring the therapeutic accessibility of genetically supported circulating proteins. Proteins uniquely measurable on the SomaScan 7k platform contribute to MR-significant associations and expand the genetically supported target space beyond proteins represented in earlier platforms. **b.** All proteins tested (UniProt, n = 1,548. For 1 UniProt, secretion pathway could not be defined). Distribution of proteins interrogated by MR, including those uniquely captured by the SomaScan 7k expansion. **c.** MR-significant proteins (UniProt, n = 574). Proteins with significant association to at least one phenotype after Bonferroni correction. **d.** Drug-targeted proteins (UniProt, n = 74). Subset of MR-significant proteins that are targets of ≥1 approved drug. **e.** Repurposing candidates (UniProt, n = 43). Subset of MR-significant proteins that are targets of approved drugs but associated with phenotypes distinct from their original therapeutic indication, indicating potential repurposing opportunities. (**f**) Proposed mechanism linking plasma FN3K levels to HbA1c as an example of genetically anchored clinical insights allowed from SomaLogic 7k measurements in plasma. Fructosamine-3-kinase (FN3K) is an intracellular protein newly quantified in plasma using the SomaLogic 7k assay. We identified a *cis*-pQTL (lead variant 17:80690014:A:G, P-value < 10^-440^) that colocalizes with the FN3K whole-blood eQTL (PP.H4 = 0.99), consistent with a shared causal variant regulating FN3K expression in whole blood. FN3K is involved in the phosphorylation of fructosamine groups bound to hemoglobin within erythrocytes (RBC, red blood cell), which leads to spontaneous hemoglobin deglycation, and thus lower glycated hemoglobin level and lower apparent diabetes risk. We hypothesize that circulating FN3K reflects abundance through vesicle release during erythrocyte aging and turnover, potentially contributing to a glycation gap between HbA1c and true glycaemic status. (**g**) Forest plot showing MR estimates for the association of genetically predicted plasma FN3K levels with HbA1c, type 2 diabetes, and fasting glucose. Colocalization supported a shared causal variant for HbA1c (PP.H4 = 0.993) and type 2 diabetes (PP.H4 = 0.986), but not fasting glucose (Supplementary Fig. 11). No evidence of an MR association was observed for fasting glucose.

Among the 7,591 significant triplets, 367 involved genes that are already targets of approved drugs but associated with diseases or traits distinct from their original exact indications, suggesting potential opportunities for drug repurposing (**Methods and Supplementary Table 7**). To prioritise findings supported by convergent molecular evidence, we evaluated colocalization with *cis*-eQTLs for the encoding gene and flagged aptamers with potential epitope-binding artefacts. Among the 7,268 MR associations with available GTEx colocalization results, 4,970 (68%) colocalized with a *cis*-eQTL for the encoding gene. Of these, 4,660 (94%) were not flagged as epitope-sensitive instruments (**Figure 3a**). Several of these high-confidence associations recapitulated well-established disease mechanisms, including CFH with age-related macular degeneration, IL6R and PCSK9 with cardiovascular disease phenotypes. Additional high-confidence associations highlighted biologically coherent disease pathways supported by tissue-relevant *cis*-eQTL colocalization, including TLR3 with hypothyroidism together with thyroid *cis*-eQTL colocalization, and MMP12 with peripheral vascular disease, aneurysmal disease, and chronic lung disease phenotypes together with colocalization in spleen, a macrophage-rich immune tissue consistent with the known role of MMP12 in inflammatory extracellular matrix remodelling^21,22^ (see **Supplementary Table 7** and phenotype landscape in **Figure 3f–g**).

Intracellular proteins comprised 48.1% (745/1,548) of all MR protein exposures (UniProt) with a cellular component annotation. These proteins were progressively depleted across the translational cascade, representing 45.8% (263/574) of proteins with significant MR associations, but only 20.3% (15/74) of MR-significant proteins with an approved drug and 18.6% (8/43) of MR-significant proteins corresponding to a potential repurposing opportunity. By contrast, an inverse enrichment trend was observed for secreted proteins: they accounted for 34.8% (539/1,548) of all MR protein exposures, but 38.3% (220/574) of proteins with significant MR associations and 48.6% (36/74) of significant proteins targeted by a drug (**Figure 3b–e**). These enrichment and depletion patterns suggest that the current therapeutic landscape remains strongly enriched for genetically supported circulating and secreted proteins, whereas intracellular proteins remain comparatively underrepresented in existing drug development pipelines.

Proteins newly measured on the SomaScan 7k platform accounted for 16% (250/1,548) of all MR protein exposures and 14% (80/574) of MR-significant proteins, demonstrating non-redundant expansion of the causal discovery space. Newly captured proteins contributed fewer drug-targeted (2/74) and repurposing (2/43) proteins compared to all proteins tested (250/1,548), suggesting that plasma proteome expansion primarily reveals genetically-supported disease mechanisms not yet represented in late-stage pharma pipelines, rather than simply reinforcing established targets (**Figure 3b–e**). Importantly, newly measured proteins on the SomaScan 7k platform, including intracellular and membrane-associated proteins, nevertheless showed coherent genetically supported disease associations. For example, genetically predicted higher levels of the intracellular mitochondrial protein citramalyl-CoA lyase (CLYBL) were associated with lower risk of vitamin B-complex deficiency (β = −0.24 per genetically predicted standard deviation increase in circulating CLYBL levels, SE = 0.023, P-value = 9.8 × 10⁻²⁷, PP.H4 = 0.999), with the *cis*-pQTL colocalizing with *CLYBL cis*-eQTLs in liver, adipose, skeletal muscle and whole blood. The direction of effect is consistent with human genetic and biochemical studies implicating *CLYBL* in mitochondrial vitamin B12 metabolism and protection from vitamin B12 depletion.^23,24^ We also found inverse associations for genetically predicted higher levels of the intracellular protein fructosamine-3-kinase (FN3K) with glycated hemoglobin (HbA1c) levels (β = -0.13, SE = 0.007, P-value = 9.62 x 10^-91^) and risk of type 2 diabetes (β = -0.05, SE = 0.007 P-value = 9.77 x 10^-12^), but not for other glycemic traits including fasting glucose or random glucose (P-value > 0.05, **Supplementary Table 7**). The lead *cis*-pQTL signal for FN3K, 17:80690014:A:G, which was also identified by a smaller mass-spectometry-based proteomic study,^25^ colocalises (PP.H4>0.98) with associations for HbA1c and type 2 diabetes, as well as with a *cis*-eQTL for *FN3K* in whole blood in GTEx, suggesting a regulatory effect of the variant. FN3K is a newly measured protein in the SomaScan 7k assay and is responsible for phosphorylating fructosamine groups bound to intracellular proteins, which leads to spontaneous dissociation of the glucose from the protein. This deglycation is known to occur on hemoglobin in erythrocytes^26,27^ leading to suggestions that variable *FN3K* activity between individuals may contribute to a ‘glycation gap’,^28^ where HbA1c levels become a less accurate measure of true glucose exposure and thus of diabetes risk or diagnosis (**Figure 3f-g and Supplementary Figure 11**). Our findings support suggestions that future efforts to account for variation in *HBB*, *G6PD* and *PIEZO1*^29,30^ when ascertaining glycemic status should also be extended to *FN3K*.

Together, these examples demonstrate that proteome expansion broadens genetically anchored disease mapping toward intracellular and membrane-associated biology that was underrepresented in earlier plasma proteomic studies but can still be of clinical significance.

## Discussion

By profiling 7,144 aptamers in 13,445 European participants using the expanded SomaScan 7k platform, we provide the most systematic evaluation to date of what proteomic expansion delivers for proteogenomic discovery science. By integrating genetic mapping, colocalization, Mendelian randomization, and detailed assay characterization, we show that increasing proteomic coverage does not simply increase the number of detectable pQTLs: rather, it extends genetically accessible biology beyond the classical secretome while revealing biological constraints that limit discovery and complicate causal inference. These findings have practical implications for the design of future proteogenomic studies and for the interpretation of pQTL-based causal analyses as platform breadth continues to expand. To facilitate future studies, we make all summary statistics publicly available through an interactive browser and download portal https://pgwas-chris-interval.gm.eurac.edu/.

A major focus of our analysis was quantifying how expansion of the plasma proteome alters the spectrum of genetically accessible biology. We assessed its consequences for *cis*-pQTL detectability and discovery rates, by systematically comparing newly measurable proteins with aptamers already present in earlier SomaScan versions. Newly added proteins had similar measurement quality metrics, were predominantly intracellular and had substantially lower expected plasma abundance. This difference in biological properties likely results from a design choice of the proteomic platform provider but may reflect a more generalizable perspective on proteomic assay expansion: as well-studied plasma proteins were prioritised in earlier platform versions, future platforms will necessarily extend to proteins with lower plasma relevance. Newly added proteins were associated with lower *cis*-pQTL discovery rates (15% for newly added proteins versus 28% for proteins previously assayed), but these differences were largely explained by their biological characteristics rather than intrinsic assay performance. This indicates the lower yield of *cis*-pQTLs is an inherent consequence of expanding proteome coverage beyond the classical secretome, although reduced measurable inter-individual variation near the limits of plasma detectability may also contribute. Importantly, despite their lower abundance, 176 novel *cis* signals arose from newly measurable proteins. Thus, even if assay expansion introduces many lower-abundance aptamers with inherently lower discovery probability, it still enables the identification of genetic associations for proteins not represented in earlier proteomic platforms. Overall, plasma proteome expansion broadens genetically accessible biology beyond the classical secretome, but increasingly encounters biological constraints imposed by low-abundance intracellular proteins, highlighting the need for larger cohorts and tissue-specific proteogenomic studies.

Colocalization analyses showed extensive sharing of genetic regulation across the circulating proteome. A central insight of this study is the characterization of the structure of *trans*-regulatory genomic hotspots, which accounted for 68% of all *trans* associations, reflecting variants with unusually broad regulatory effects on the circulating proteome. These hotspots included loci with known pleiotropic effects and loci that may represent novel shared regulatory pathways. We noted that some hotspots may represent signals related to technical or sample handling variation, emphasizing the importance of such analyses. Using colocalization patterns across proteins, we characterized these regions at finer resolution. Some hotspots, such as the *GCKR* locus, were consistent with a single highly pleiotropic driver, whereas others, such as the chromosome 3 FETUB/HRG/KNG1 region, resolved into several neighbouring clusters driven by distinct variants. These findings demonstrate that extreme *trans* pleiotropy can arise from both single and multiple underlying signals within the same genomic region, and that such structures become visible only when large numbers of proteins are jointly quantified. We also observed extensive multi-target regulation in *cis*: 105 pairs of *cis*–*cis* colocalizing signals involved clusters of related proteins, as well as pairs without obvious functional relatedness. As proteomic breadth increases, these multi-target *cis* effects become increasingly apparent, underscoring that *cis*-pQTLs should not be assumed to uniquely tag a single protein. This growing complexity highlights the need for integrative approaches when using pQTLs to infer causal mechanisms.

Integration with GTEx *cis*-eQTLs revealed tissue-specific enrichment in tissues directly relevant to protein secretion into plasma (e.g., blood, adipose, and muscle) while also highlighting that pQTLs frequently regulate multiple genes, complicating causal gene assignment. Importantly, colocalization identifies shared association signals, rather than causal genes, and is not intended to uniquely assign mechanistic responsibility within complex loci. As proteomic datasets expand, these shared architectures will only become more pronounced, exposing additional layers of coordinated regulation but also increasing ambiguity in causal assignment. This underscores the need for integrative frameworks that combine colocalization with context-specific MR, tissue-, organ- and disease mechanism-relevant mediation analyses, and experimental follow-up.

Using *cis*-pQTL signals as genetic instruments, we identified 6,340 protein–phenotype pairs with potential causal roles across 2,003 diseases and traits, including potential drug repurposing opportunities. Proteome expansion increases the scope for genetically-anchored causal inference, extending drug repurposing analyses to proteins not represented in earlier platforms, even if most drug-target validation and repurposing signals still arise from proteins present in earlier 5k assays. Notably, secreted proteins were overrepresented in MR associations and even more in drug-target validation and repurposing. This pattern may reflect both biological and technical factors: secreted proteins are more directly accessible as circulating targets and have also been historically more studied^31^ and therefore might be more frequently represented in existing drug development pipelines, while their higher circulating concentrations likely increase statistical power for Mendelian randomization analyses. Overall, our results indicate that proteome expansion extends the space of genetically supported protein–phenotype associations, but that, when pQTL are studied in plasma, the immediate translational signal remains concentrated among well-characterized, secreted proteins.

This work illustrates the utility of proteogenomic analyses in refining the biological understanding of circulating proteins and informing therapeutic target prioritization. However, several limitations should be considered. The restriction to participants of European ancestry limits generalizability, particularly for *trans*-pQTL hotspot findings where between-study heterogeneity was already substantial within European cohorts; meta-analysis across two independent cohorts nonetheless provided a degree of internal replication that likely reduced false-positive findings. Aptamer cross-reactivity and epitope-sensitive effects, particularly at loci involving protein-altering variants (PAVs), represent an important limitation of affinity-based proteomic assays. While some PAV-associated pQTLs likely reflect genuine biological effects on protein abundance, secretion, stability, or processing, others may arise from altered aptamer binding affinity rather than quantitative differences in circulating protein levels. These mechanisms cannot be fully distinguished using genetic association data alone. To mitigate this issue, we systematically annotated potentially epitope-sensitive signals and integrated orthogonal evidence, including *cis*-eQTL colocalization, when prioritizing downstream Mendelian randomization findings. Nevertheless, PAV-linked pQTLs should be interpreted cautiously, particularly when used as instruments for causal inference, and future studies integrating orthogonal proteomic technologies and functional validation will be important to refine their interpretation. Last, MR analyses rely on assumptions, including the absence of horizontal pleiotropy, that may not always hold, underlining the need for triangulation with orthogonal approaches and functional follow-up studies.

For novel biology uncovered, future work should focus on replication and fine-mapping in diverse ancestries, integration with additional molecular layers, and experimental follow-up to assess mechanisms underlying identified associations. Combining MR with mediation analyses and functional perturbation studies will further strengthen clinical causal inference and advance translation into therapeutic development. Replication of pQTLs in relevant tissues will be essential for intracellular proteins, and the biological scope of upcoming expanded proteomic platforms should be carefully considered when designing future population-scale pQTL studies.

In summary, this study demonstrates the value of large-scale proteogenomics for mapping the regulatory architecture of the plasma proteome and identifying disease-relevant associations, while revealing that proteome expansion extends genetically accessible biology at the cost of increasing biological and statistical constraints, insights that should inform the design of future high-throughput proteomic platforms.

## Data Availability

GWAS summary data are available for scientific and non-commercial use on https://pgwas-chris-interval.gm.eurac.edu after registration and statement of data usage purpose. We currently provide two resources: (1) A PheWeb that can be used to visualise and browse the results, (2) a download page to download all 7,144 summary statistics. To eliminate the possibility of reidentification of study participants, the precision of MAF, beta, SE and P-values were reduced.

## Authors contributions

Study conceptualization: E.D.A, C.F., A.S.B. Project coordination: S.C, C.G., A.M.. Design and funding of contributing cohorts: CHRIS: C.P, P.P.P., C.F., F.S.D., J.R., M.G., R.L., D.M., R.B., H.B., A.D.G., C.E., E.D.A.; INTERVAL: E.D.A, A.S.B, J.D.. Design of the analyses and drafting of the manuscript: S.C., C.G., A.S.B., A.M., E.K., D.G-S., G.P., M.F.. Proteomic, phenotypic, genotype acquisition and QC: CHRIS: E.K., M.F., S.C., C.F., C.P., F.S.D., J.R., M.G., R.L., D.M., R.B., H.B., A.D.G., E.D.A, C.E.; INTERVAL A.M., X.J., S.C., G.C., C.G., E.D.A., A.S.B., J.D. Cohort genetic associations analyses, sensitivity analysis, harmonization and meta-analysis: M.F, S.C, C.G., G.P., G.C. Regional associations, conditional analyses and fine-mapping: D.G.-S., C.G., S.C. pQTL colocalization: D.G.-S., G.P., A.M., C.G., S.C. eQTL colocalization: E.K. Mendelian Randomization and phenotype colocalization: A.P., B.F., G.C., K.G.-P., G.P., C.G., D.G.-S.. Variants, aptamers and signals annotation: S.C., M.F., D.G.-S., A.M., G.P., X.Y. E.K., M.C.M. Development of the interactive data portal: E.K., H.B.. All authors contributed to the interpretation of the results and critically reviewed the manuscript.

## Acknowledgment

The CHRIS study was funded by the Autonomous Province of Bolzano/Bozen - South Tyrol - Department of Innovation, Research, University and Museums and supported by the European Regional Development Fund (FESR1157). CHRIS SomaLogic proteomic measurements were funded by Fondazione Human Technopole.

The CHRIS study thanks all study participants, the Healthcare System of the Autonomous Province of Bolzano Bozen - South Tyrol, and all Eurac Research staff involved in the study (https://www.eurac.edu/chrisack). Bioresource Impact Factor Code: BRIF6107.

Participants in the INTERVAL randomized controlled trial were recruited with the active collaboration of NHSBT England (https://www.nhsbt.nhs.uk/), which has supported fieldwork and other elements of the trial. DNA extraction and genotyping were co-funded by the National Institute for Health and Care Research (NIHR), the NIHR BioResource (https://bioresource.nihr.ac.uk/) and the NIHR Cambridge Biomedical Research Centre (BRC-1215-20014). Proteomic measurements were funded by Fondazione Human Technopole. The academic coordinating centre for the INTERVAL study was supported by core funding from the: NIHR Blood and Transplant Research Unit (BTRU) in Donor Health and Genomics (NIHR BTRU-2014-10024), NIHR BTRU in Donor Health and Behaviour (NIHR203337), UK Medical Research Council (MR/L003120/1), British Heart Foundation (SP/09/002; RG/13/13/30194;

RG/18/13/33946 and RG/F/23/110103), BHF Chair Award (CH/12/2/29428) and NIHR Cambridge BRC (BRC-1215-20014; NIHR203312). The academic coordinating center thanks blood donor center staff and blood donors for participating in the INTERVAL study. This work was supported by Health Data Research UK, which is funded by the UK Medical Research Council, Engineering and Physical Sciences Research Council, Economic and Social Research Council, Department of Health and Social Care (England), Chief Scientist Office of the Scottish Government Health and Social Care Directorates, Health and Social Care Research and Development Division (Welsh Government), Public Health Agency (Northern Ireland), British Heart Foundation and Wellcome. The views expressed are those of the authors and not necessarily those of the NIHR or the Department of Health and Social Care.

This research is based in part on data and summary statistics from the Million Veteran Program, Office of Research and Development, Veterans Health Administration, and was supported by award no. MVP000. Selected analyses used resources from the Knowledge Discovery Infrastructure at Oak Ridge National Laboratory, supported by the Office of Science of the U.S. Department of Energy under Contract No. DE-AC05-00OR22725 and the Department of Veterans Affairs Office of Information Technology Inter-Agency Agreement with the Department of Energy under IAA No. VA118-16-M-1062. This publication does not represent the views of the Department of Veterans Affairs or the United States Government.

We acknowledge the support of Edoardo Giacopuzzi for the use of the GWAS regenie pipeline for INTERVAL.

## Ethics statement

The Ethics Committee of the Healthcare System of the Autonomous Province of Bolzano/Bozen - South Tyrol approved the CHRIS baseline protocol on 19 April 2011 (21/2011) with updates approved on 12 October 2015, 28 October 2015, and 28 January 2016 and the CHRIS proteomic protocol on 23 July 2023. The study conforms to the Declaration of Helsinki, and with national and institutional legal and ethical requirements.

The INTERVAL study was approved by the UK National Research Ethics Service (11/EE/0538) and all participants gave informed consent.

Analyses involving Million Veteran Program (MVP) data were conducted in accordance with the policies and approvals of the Veterans Health Administration. All MVP participants provided informed consent, and study protocols were approved by the appropriate institutional review boards and research oversight committees.

## Online Methods

### Study cohorts

We analyzed plasma proteomic and genomic data from two European-ancestry population studies: INTERVAL and the Cooperative Health Research in South Tyrol (CHRIS) study.

The INTERVAL^8^ study is a randomized pragmatic trial of blood donation frequency conducted across 25 National Health Service Blood and Transplant centers in England. Participants were recruited between 2012 and 2014, provided written informed consent, and were approved by the National Research Ethics Service (11/EE/0538). Eligible participants were aged 18–80 years and were generally healthy at recruitment, as individuals with major diseases or recent illness were ineligible for blood donation. Non-fasted blood samples were collected at the beginning of routine donation visits, and participants completed detailed questionnaires on lifestyle, demographics, anthropometry, and diet. A subset of 9,679 participants was selected for SomaScan proteomic profiling using a case–cohort design, and 9,251 participants of European ancestry with both proteomic and genomic data were included in the present analyses.

The CHRIS^9,10^ study is a population-based cohort conducted in the Vinschgau/Venosta district in South Tyrol, Italy, designed to investigate the genetic and molecular basis of age-related conditions. Between 2011 and 2018, 13,393 adult residents (≥18 years) were recruited. Following overnight fasting, participants underwent blood and urine collection, anthropometric and clinical measurements, and standardized interviews assessing health status and lifestyle exposures. SomaScan proteomic profiling was performed in 4,229 participants selected to maximize overlap with other molecular datasets, and 4,194 participants of European ancestry with high-quality genomic and proteomic data were included in this study.

Additional cohort details are provided in the Supplementary Methods (**SM1**).

### Protein quantification

Plasma protein levels were measured using the SomaScan 7k (v4.1) aptamer-based proteomic assay (SomaScan)^32^, which quantifies 7,289 aptamers targeting 6,381 proteins. In INTERVAL, assays were performed in two batches; in CHRIS, assays were performed in a single batch using the same protocol. SomaScan performed standard within-run and between-run normalization using hybridization controls and calibrator samples.

Quality control procedures included assessment of assay precision using coefficients of variation (CVs) derived from technical replicates and estimation of per-aptamer limits of detection (LOD) using buffer samples. Aptamers with poor detectability were excluded based on predefined thresholds. After quality control, measurements for 7,145 aptamers were retained in INTERVAL and 7,281 aptamers in CHRIS, with a harmonized set of 7,144 aptamers used for downstream analyses. Protein abundances were log-transformed and inverse-rank normalized prior to genetic association testing. Detailed assay processing, quality control metrics, and batch comparability analyses are described in the Supplementary Methods (**SM2**).

### Genotyping, imputation, and ancestry inference

INTERVAL participants were genotyped using the ThermoFisher UK Biobank Axiom array, and genotypes were imputed using the 1000 Genomes Phase 3–UK10K reference panel.^33^ CHRIS participants were genotyped on the Illumina HumanOmniExpressExome and Omni2.5Exome arrays, with imputation performed using the Haplotype Reference Consortium reference panel. Standard quality control procedures were applied in both cohorts, including filtering on sample and variant call rates, Hardy–Weinberg equilibrium, minor allele count (MAC ≥ 10), and imputation quality (INFO > 0.7).

Within each cohort, variants were restricted to autosomes and multi-allelic variants were split and normalized. Population structure was assessed using principal component analysis, and individuals identified as ancestry outliers were excluded.

To ensure a consistent definition of effect alleles prior to association testing, alleles were reordered alphabetically (chr:pos:effect allele:non-effect allele) and effect sizes were defined relative to this standardized allele ordering.

Full details of genotype processing, imputation, and ancestry inference are provided in the Supplementary Methods (**SM3**).

### Genome-wide association analyses

Genome-wide association studies (GWAS) of plasma protein levels were performed separately in INTERVAL and CHRIS using REGENIE^34^ (v3.3), which implements a two-step whole-genome regression framework to account for population structure and relatedness. In the first step, polygenic predictions were estimated using genotyped variants under a leave-one-chromosome-out scheme. In the second step, single-variant association testing was performed using linear regression, conditional on the polygenic predictions.

Based on analyses of technical factors associated with aptamer variability (**SM2**), association models in INTERVAL included batch, age, sex, season of blood collection, time from blood draw to processing, and the first ten genetic principal components, and were applied to 9,251 samples. In CHRIS, the same covariates were used except batch, and models were applied to 4,194 samples. In both cohorts, residuals were computed and used as phenotypes for GWAS. Genomic inflation was assessed using the genomic control lambda statistic.

### Meta-analysis

Summary statistics from INTERVAL and CHRIS were combined using inverse-variance weighted meta-analysis implemented in METAL.^35^ Allele matching across cohorts were performed automatically by METAL. Only variants present in both cohorts were retained for downstream analyses.

### Definition of *cis* and *trans* pQTLs and identification of regional associations

Aptamers were mapped to their corresponding protein-coding genes using UniProt and Entrez Gene identifiers, and transcription start sites (TSS) were assigned based on the GRCh37 reference genome. For each aptamer, *cis* regions were defined as ±500 kb windows around the TSS of the corresponding target gene(s); associations overlapping any *cis* region were classified as *cis*, and all remaining associations were classified as *trans*.

To identify significant regional associations in the meta-analysis, variants with P-value < 1 × 10⁻⁶ were first selected and grouped into loci if they were on the same chromosome and within 3 Mb of each other. A locus was considered significant if it contained at least one variant exceeding the Bonferroni-corrected significance threshold used in this study (P-value < 1.26 × 10^−11^).^36^ Two loci were excluded from downstream analyses due to challenges for fine-mapping and interpretation: the HLA region (chr6:28,477,797–33,448,354; GRCh37) and an extended region around NLRP12 on chromosome 19 (chr19:54,300,000–54,360,000; GRCh37)^37^.

### Conditional analysis and fine-mapping

To identify conditionally independent association signals, regional loci were extended by ±100 kb and analyzed using stepwise conditional regression implemented in GCTA-COJO.^38^ Variants with joint P-values below the multiple-testing threshold were retained as independent signals. Fine-mapping was performed using approximate Bayes factors to derive 99% credible sets for each conditional signal. Full details are provided in Supplementary Methods (**SM14-15**).

### Colocalization analyses

Colocalization^39^ analyses were performed to identify shared causal variants across pQTL signals and between pQTLs and expression quantitative trait loci (eQTLs). For pQTL–pQTL colocalization, tests were restricted to pairs of conditionally independent pQTL signals whose 99% credible sets shared at least one variant. Bayesian colocalization was performed using coloc, and colocalization was inferred when the posterior probability for a shared causal variant exceeded 0.8.

For pQTL–eQTL colocalization, *cis*-eQTL summary statistics from GTEx^17^ v8 across 49 tissues were used, and colocalization was performed within the boundaries of the pQTL regional associations using conditional pQTL summary statistics. Tissue-specific enrichment of colocalization was assessed by comparing observed and expected numbers of colocalized signals. Additional methodological details are provided in the Supplementary Methods (**SM15**).

### Mendelian randomization

Two-sample Mendelian randomization (MR) analyses were performed using *cis*-pQTLs as instruments to evaluate potential causal effects of circulating proteins on diseases and complex traits. Instruments were selected from conditionally independent *cis* signals and filtered to exclude weak instruments (F<10). Outcome summary statistics were obtained from large-scale GWAS meta-analyses from the Million Veteran Program,^18^ UK Biobank,^19^ and FinnGen,^20^ encompassing 2,003 harmonized phenotypes.^6^

MR analyses were conducted using the Wald ratio for single-instrument exposures and inverse-variance weighted methods for multi-instrument exposures. Statistical significance was assessed using a Bonferroni correction across all MR tests performed, defined at the level of tested SeqID–gene–phenotype triplets (0.05 / 3,497,303). Drug repurposing analyses were conducted by integrating MR results with curated drug–target–indication mappings. For MR-significant aptamer–gene–phenotype triplets, we additionally performed Bayesian colocalization between *cis*-pQTL summary statistics and the corresponding outcome GWAS signals using the coloc package,^39^ following the general framework described in Ferolito et al.^6^ Colocalization was performed using marginal *cis*-pQTL and outcome GWAS summary statistics within a ±250-kb window around each MR instrument, retaining variants with MAF > 1%. Strong evidence of shared protein and phenotype association signals was defined as posterior probability for hypothesis 4 (PP.H4.abf) > 0.8. Additional details are provided in the Supplementary Methods (**SM19**).

## Supplementary Figures

**Figure S1.**
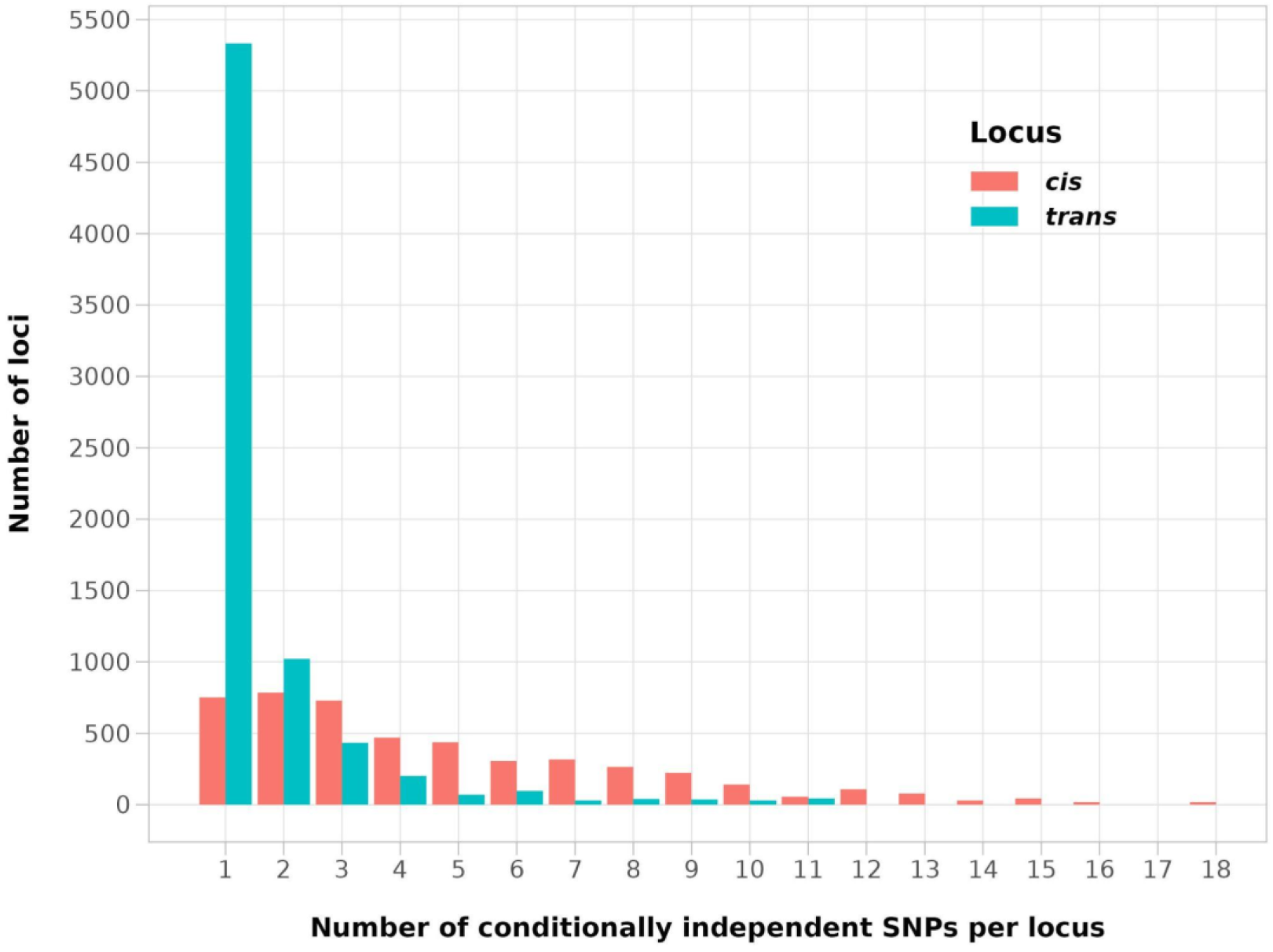
Number of conditionally independent variants per regional association.

**Figure S2.**
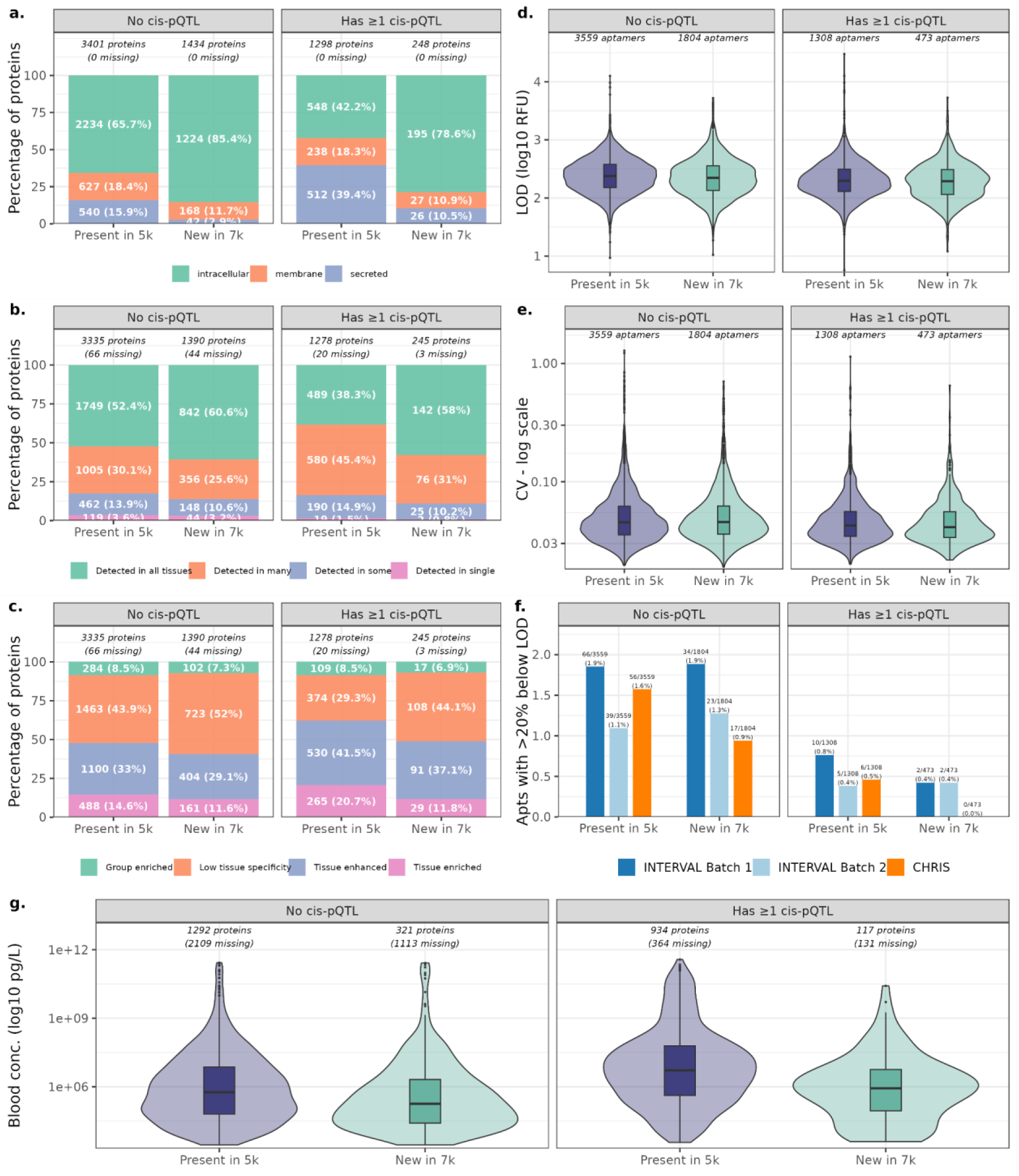
Characteristics of proteins and aptamers stratified by *cis*-pQTL status in the SomaScan 7k assay. Proteins and aptamers are stratified by *cis*-pQTL status and by assay version, indicating whether the protein was previously measured in the SomaScan 5k (v4.0) platform (“Present in 5k”) or newly introduced in the SomaScan 7k (v4.1) platform (“New in 7k”). For protein-level panels, *cis*-pQTL status was defined at the UniProt level based on whether at least one quality-controlled aptamer targeting that protein had a significant *cis-*pQTL (±500 kb of the start of the gene encoding the target protein) so that each protein contributes to a single *cis*-pQTL category. (**a–c**) Distribution of biological annotations among proteins with and without *cis*-pQTLs, shown for protein subcellular localization (a), RNA tissue distribution (b), and RNA tissue specificity (c). Analyses were performed at the protein level, with one record per UniProt identifier. Proteins with ambiguous or missing gene-based annotations, including multi-gene mappings, were treated as missing for the relevant panel and excluded from percentage calculations. For panel **b**, Human Protein Atlas RNA tissue distribution categories are defined as detected in all tissues, detected in many tissues (at least one third but not all tissues), detected in some tissues (more than one but fewer than one third of tissues), or detected in a single tissue. For panel **c**, “group enriched” denotes genes with at least four-fold higher average mRNA expression in a group of 2–5 tissues compared with all other tissues. (**d–e**) Distributions of assay detectability and variability summarized at the aptamer level. Shown are the mean log_−10_-transformed limit of detection (LOD; d) and the mean coefficient of variation (CV; e), averaged across available measurement batches. Only QC-passing aptamers with non-missing values for the corresponding metric were included. Numbers indicate the number of aptamers in each group. (**f**) Proportion of QC-passing aptamers with a mean proportion of samples below the LOD exceeding 20% in each batch (INTERVAL batch 1, INTERVAL batch 2, and CHRIS), stratified by *cis*-pQTL status and assay version. Missing batch-specific percentages were treated as 0. (**g**) Distribution of expected plasma protein concentrations (log_−10_ pg/L), obtained from the Human Protein Atlas, stratified by *cis*-pQTL status and assay version membership. For panel g, analyses were performed at the protein level; numbers indicate proteins with available concentration data and missing values.

**Figure S3.**
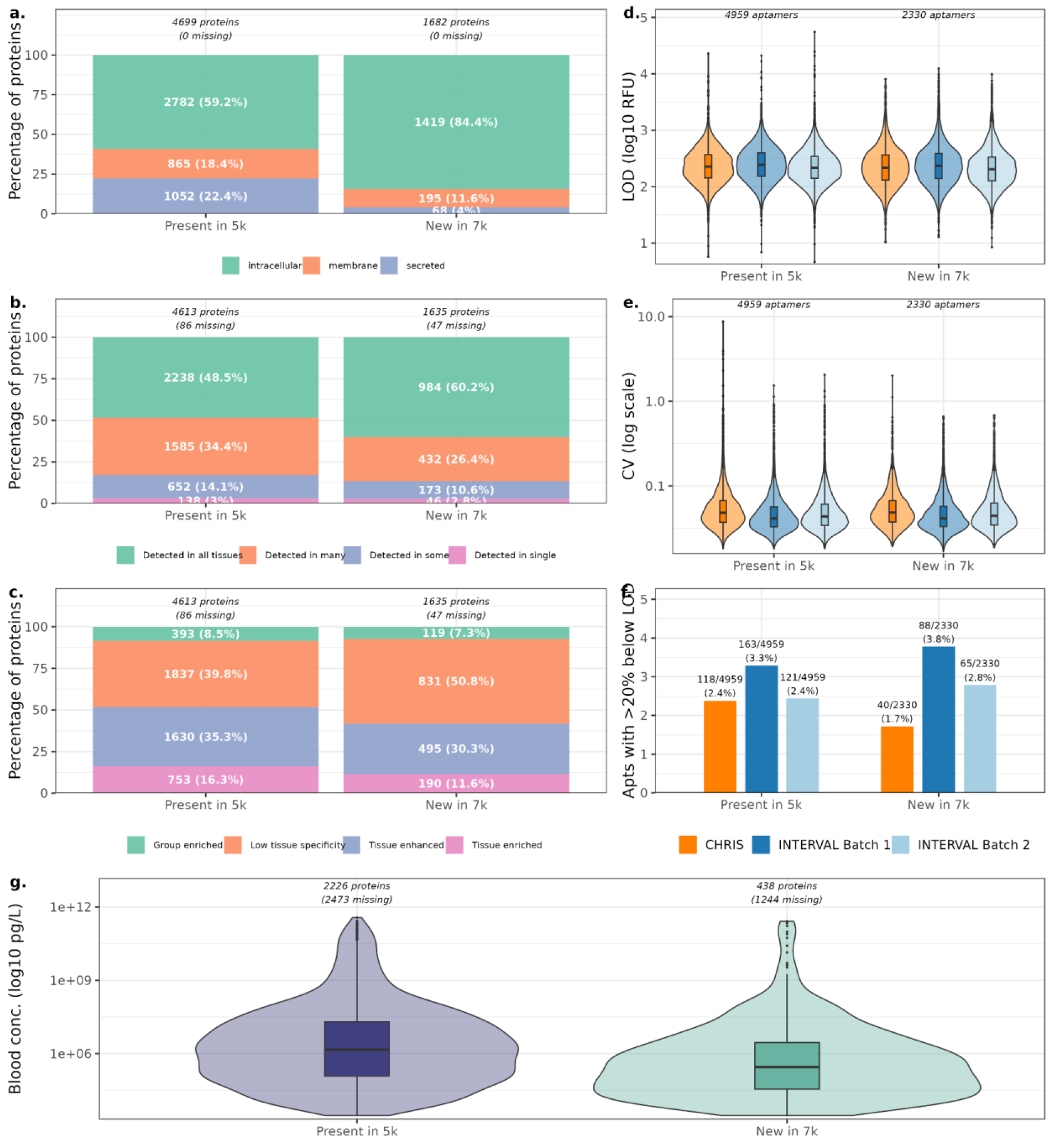
Overview of the proteins covered by SomaScan 7k versus Somascan 5k. Before LOD filtering, the dataset contained 7,289 aptamers mapping to 6,381 proteins, of which 1,682 proteins (26.4%) were newly added in the 7k assay and not present in the previous 5k (v4.0) platform. **(a–c)** Distribution of biological annotations for proteins measured on the SomaScan 7k assay, stratified by whether proteins were previously assessed in the 5k platform (“Present in 5k”) or newly introduced in the 7k platform (“New in 7k”). Shown are protein subcellular localization (a), RNA tissue distribution (b), and RNA tissue specificity (c). Bars represent percentages of proteins within each category; totals above the bars indicate the number of proteins included and the number with missing annotations. For panel b, Human Protein Atlas RNA tissue distribution categories are defined as detected in all tissues, detected in many tissues (at least one third but not all tissues), detected in some tissues (more than one but fewer than one third of tissues), or detected in a single tissue. For panel c, “group enriched” denotes genes with at least four-fold higher average mRNA expression in a group of 2–5 tissues compared with all other tissues. **(d–e)** Quality control metrics for aptamers measured across three batches (INTERVAL batch 1, INTERVAL batch 2, and CHRIS). Violin plots show distributions of log10-transformed LOD (d) and CV (e); panel e is displayed on a log10 scale. Numbers indicate the number of aptamers in each group. **(f)** Proportion of aptamers with more than 20% of samples below the LOD in each batch (INTERVAL batch 1, INTERVAL batch 2, and CHRIS). **(g)** Distribution of expected plasma protein concentrations (log10 pg/L) from the Human Protein Atlas, stratified by assay version membership. Numbers indicate proteins with available concentration data and missing values.

**Figure S4.**
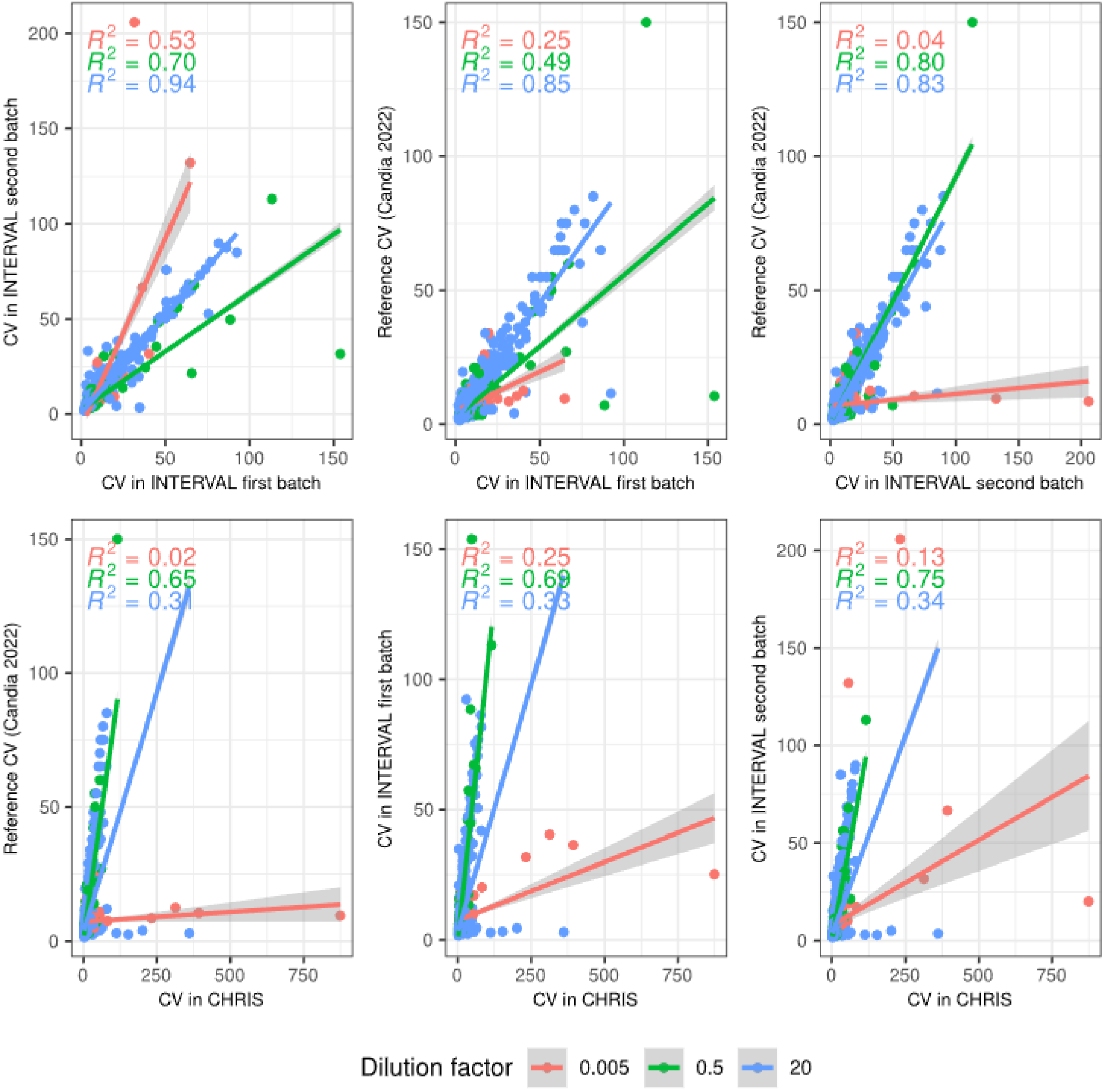
Within-batch variability. Comparisons between coefficients of variation (CVs), estimated per aptamer within INTERVAL first batch, INTERVAL second batch and CHRIS and with the literature (Candia et al., 2022). Colors represent dilution factors for the corresponding aptamer.

**Figure S5.**
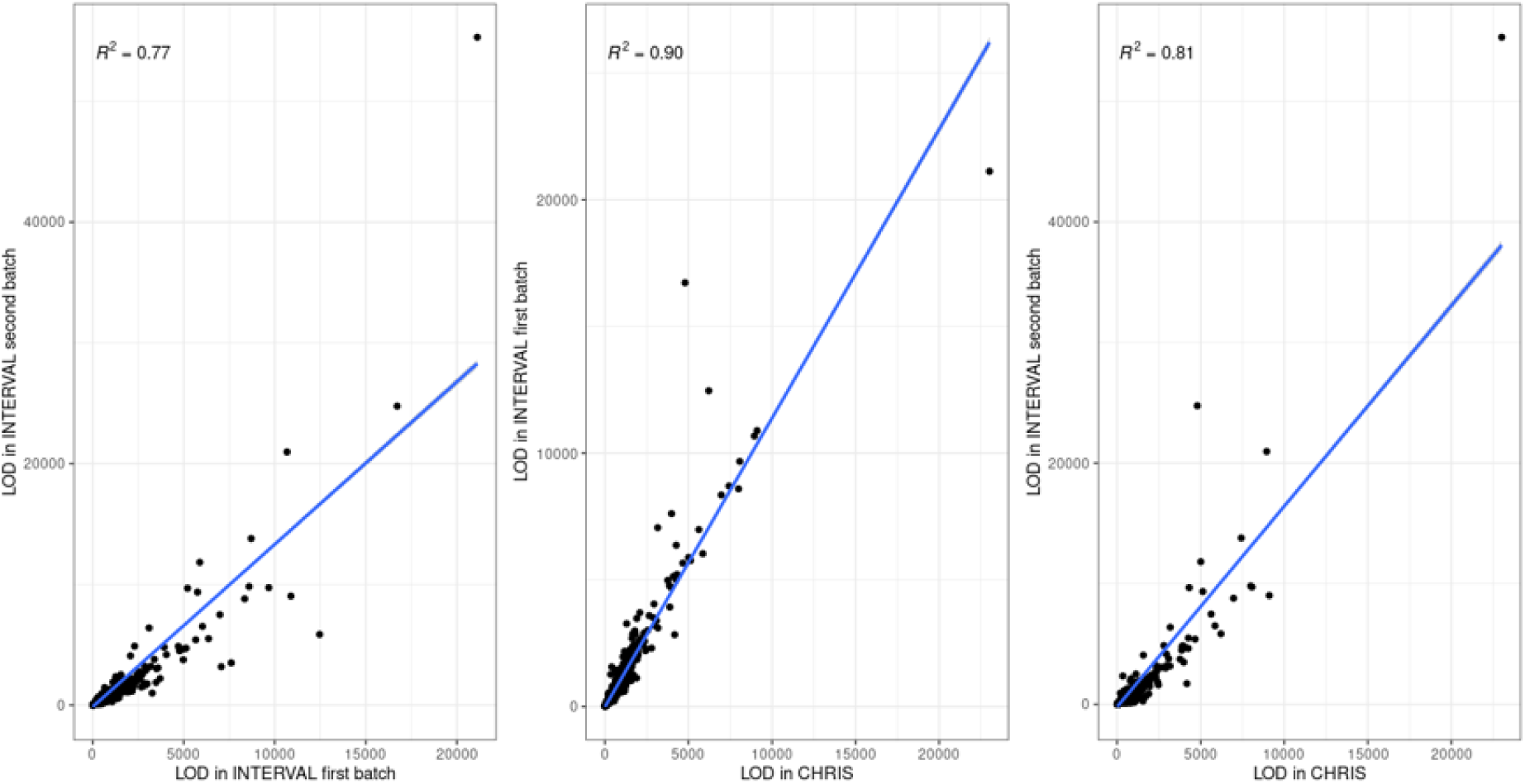
Comparisons of limits of detection across batches. Comparisons between limits of detection (LOD), estimated per aptamer using buffer samples within the two INTERVAL batches and the CHRIS study.

**Figure S6.**
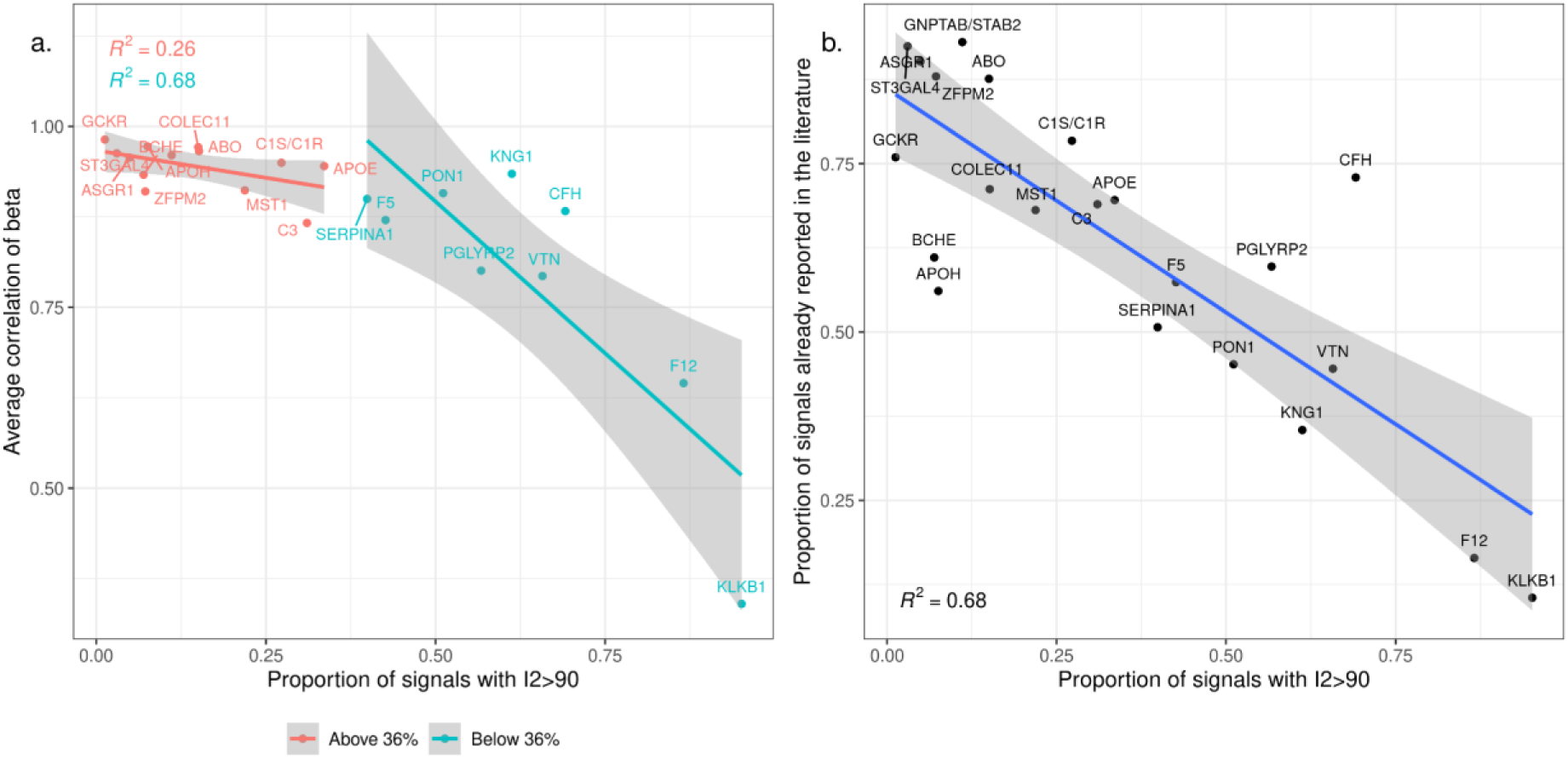
Variability of proportion of heterogeneous signals across hotspots. Proportion of heterogeneous signals (I^2^>=90 for the most significant variant in the locus) by hotspot (x-axis) according to the correlation of Beta coefficient in the 2 studies (y-axis). Color corresponds to the proportion of heterogeneous signals being higher or lower than the whole proportion among all signals (36%).

**Figure S7:**
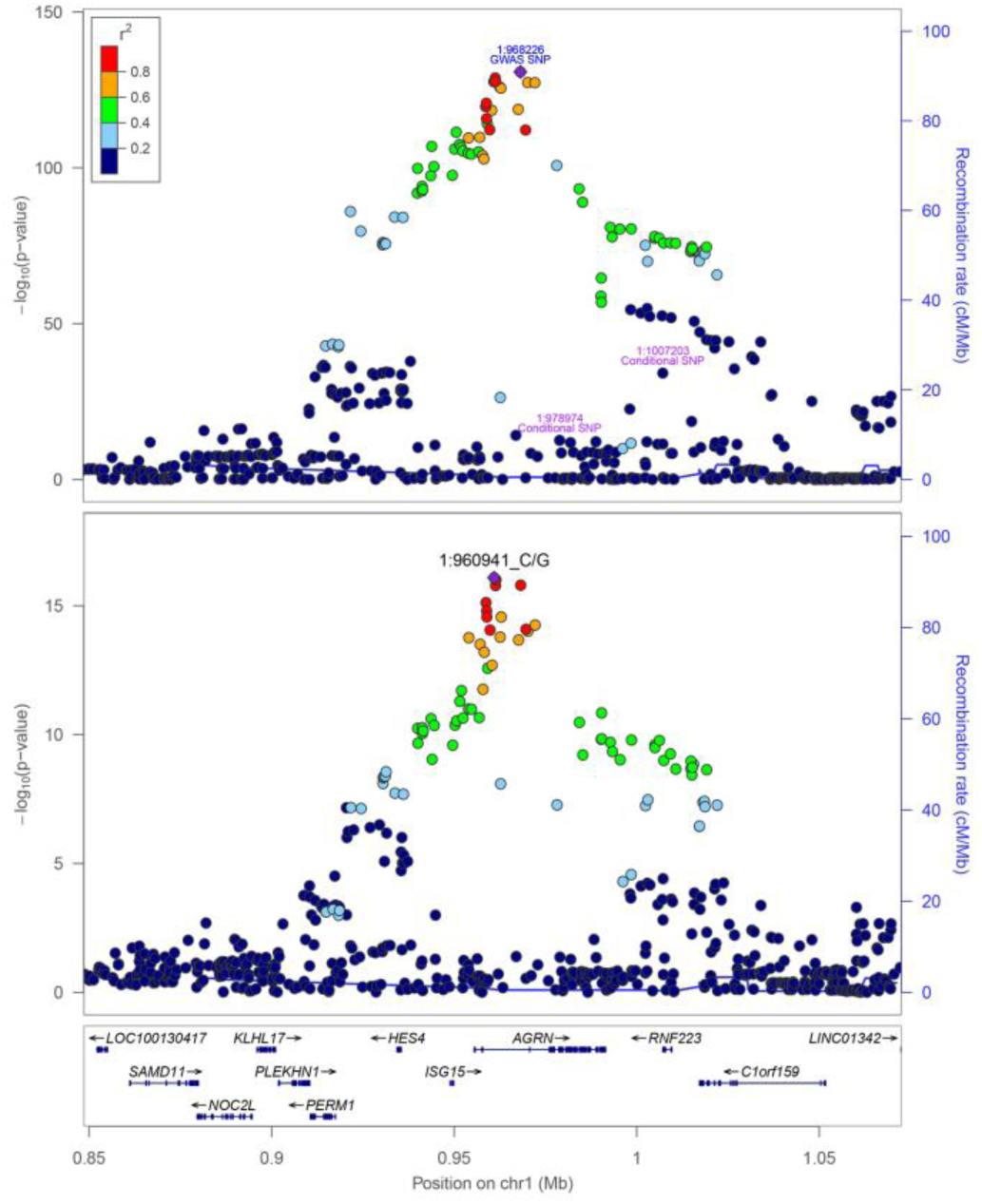
Stacked regional association plots of the two colocalized pQTLs. Bottom, LocusZoom plot of the *trans*-pQTL for *TNC* aptamer (seq.2728.62). Top, LocusZoom plot of the *cis*-pQTL for *AGRN* aptamer (seq.15483.377) where all the other conditional pQTLs were highlighted. The marginal GWAS results from meta-analysis were used to depict the pQTLs at a fixed window of *chr1:848,527-1,072,498* (GRCh37). The LD color codes computed from the INTERVAL’s genotype data. The plot was generated with LocusZoom v1.4.

**Figure S8:**
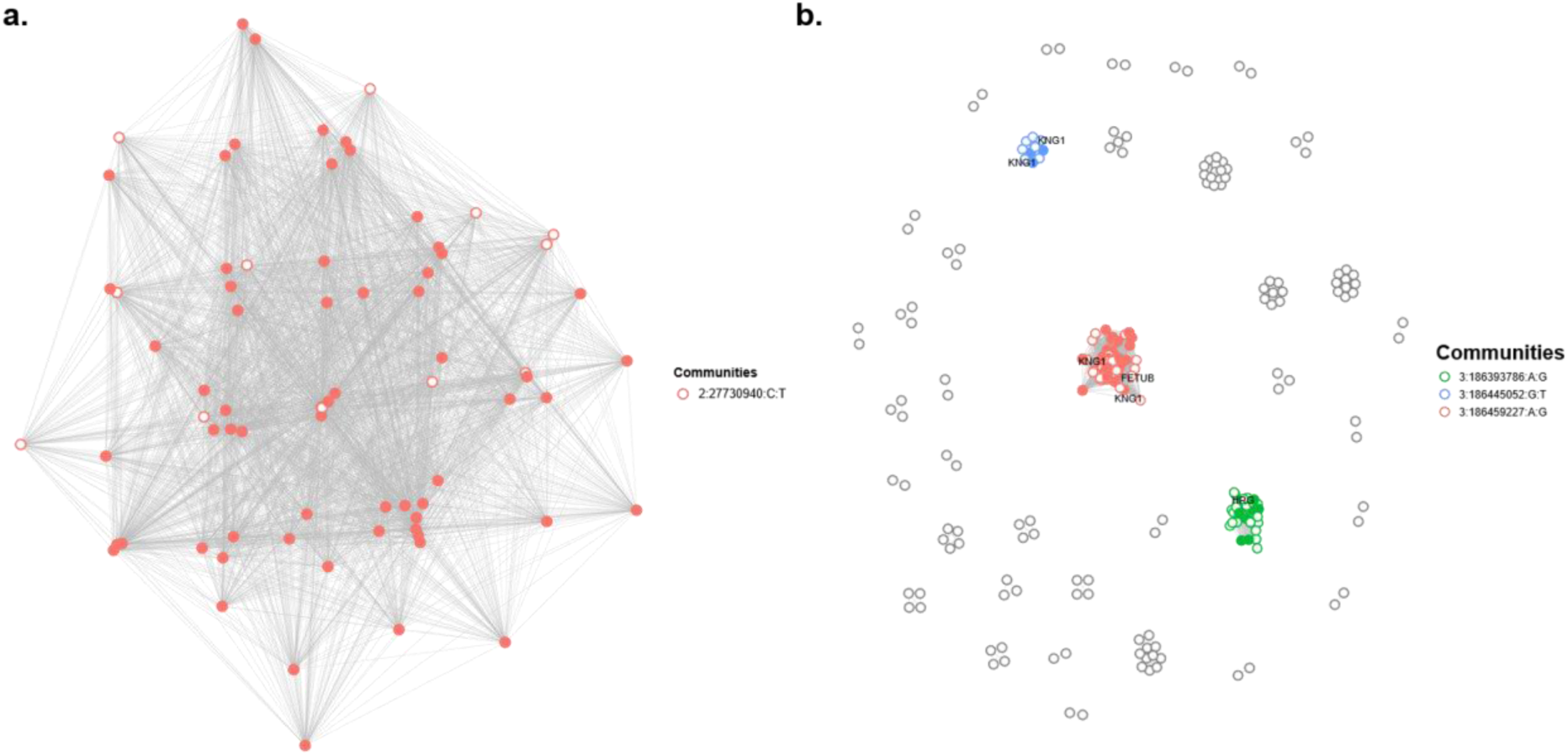
Colocalization networks for the 2 example hotspots of Figure 2. **a.** Colocalization network of an example homogeneous hotspot on chromosome 2 (GCKR locus). Each node represents a significant pQTL conditional signal, with the ones with a different index variant then rs1260326 (2:27730940:C:T) being highlighted in white. One single community has been identified. **b.** Colocalization network of an example heterogeneous hotspot on chromosome 3. Each node represents a significant pQTL conditional signal. 3 main communities have been identified. Each community is named after the index SNP of the conditional *cis*-association involved. The pQTL conditional signals not involving the corresponding SNP are highlighted in white.

**Figure S9.**
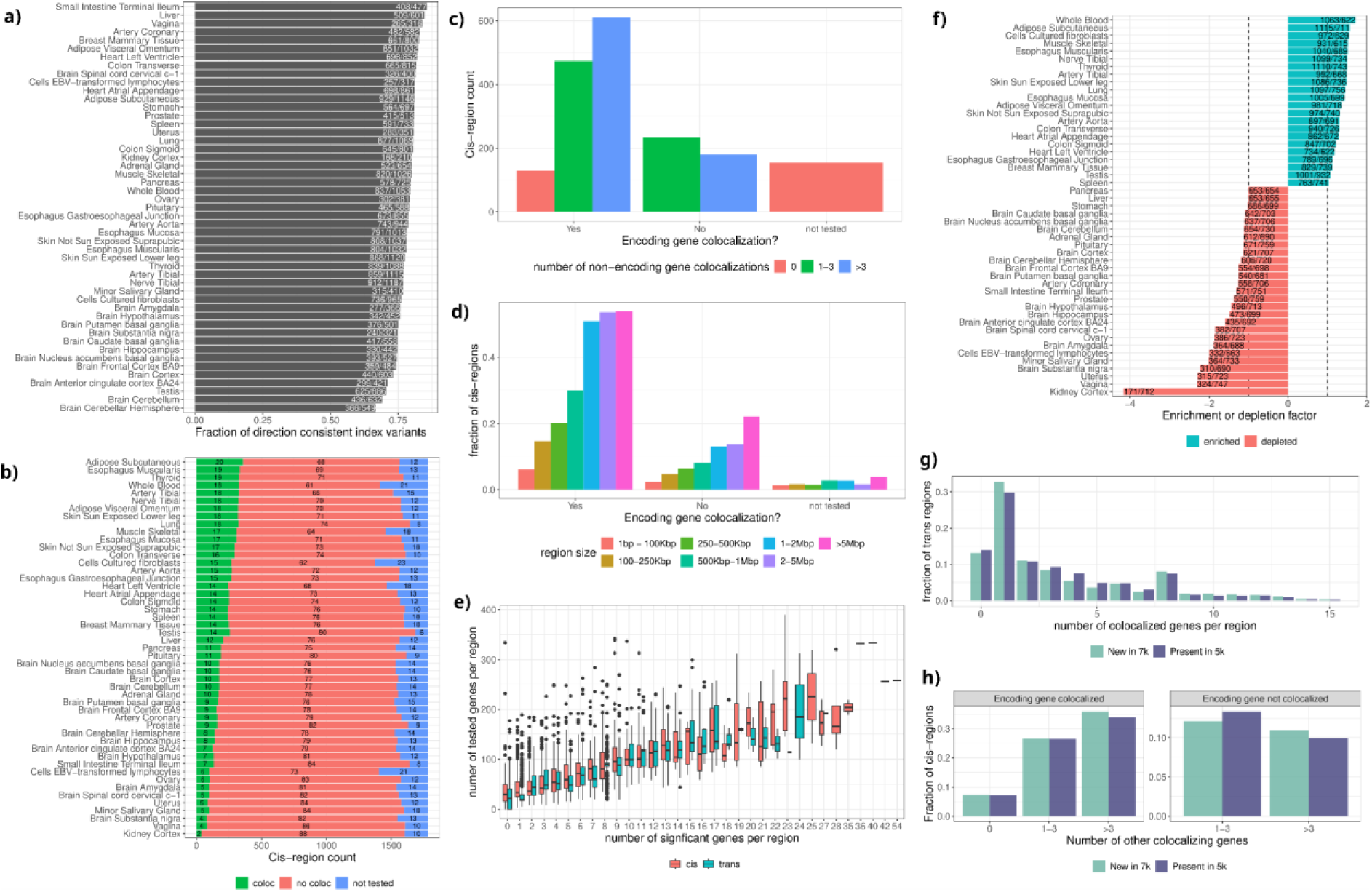
Colocalization results of meta-analysis pQTL with GTEX eQTL and eQTL lookup. (**a**) Fraction of pQTL index variants that have consistent effect direction at nominally significant (p<0.05) eQTLs. The numbers in the bar indicate the number of consistent pQTLs / the total number of nominally significant eQTLs. (**b**) Colocalization of *cis*-pQTLs with eQTLs for the gene encoding the protein across 49 GTEx tissues, highlighting tissue-specific patterns. The stacked bar plot shows, for each tissue, the number of *cis*-pQTL loci where the protein-coding gene colocalizes, does not colocalize, or was not tested due to absence in the eQTL data. (**c**) Count of *cis*-pQTL regions (n = 1,784) stratified by whether the gene encoding the protein colocalized with eQTLs in at least one tissue and the number of additional colocalizing genes within the region in at least one tissue. (**d**) Boxplot of the number of genes tested for colocalization, stratified by the number of significantly colocalizing genes found per region, for *cis*- and *trans*-pQTL loci. (**e**) Fraction of *cis*-regions with a specific size stratified by target gene colocalization. Bars of one color (one region size) sum up to 1. (**f**) Barplot of colocalization enrichment or depletion factor per tissue in *cis* regions. Values > 1 indicate an x-fold enrichment of colocalizations in the tissue, values < -1 indicate a x-fold depletion. The numbers in the bar represent the number of observed colocalizations / the number of expected colocalizations (see **Methods**). (**g**) Fraction of new and previously assessed *trans* loci regions with a specific number of colocalizing genes per locus, stratified according to the corresponding protein being newly assessed in the 7k version of SomaScan assay or notLB region. Bars of both colors sum up to 1 each, x-axis is capped at 15. (**h**) Fraction of *cis*-regions stratified by whether the target gene is colocalized with eQTLs in at least one tissue, the number of colocalizing genes other than the target gene encoding the protein and whether the protein is new in the 7k version platform or was previously assessed.

**Figure S10.**
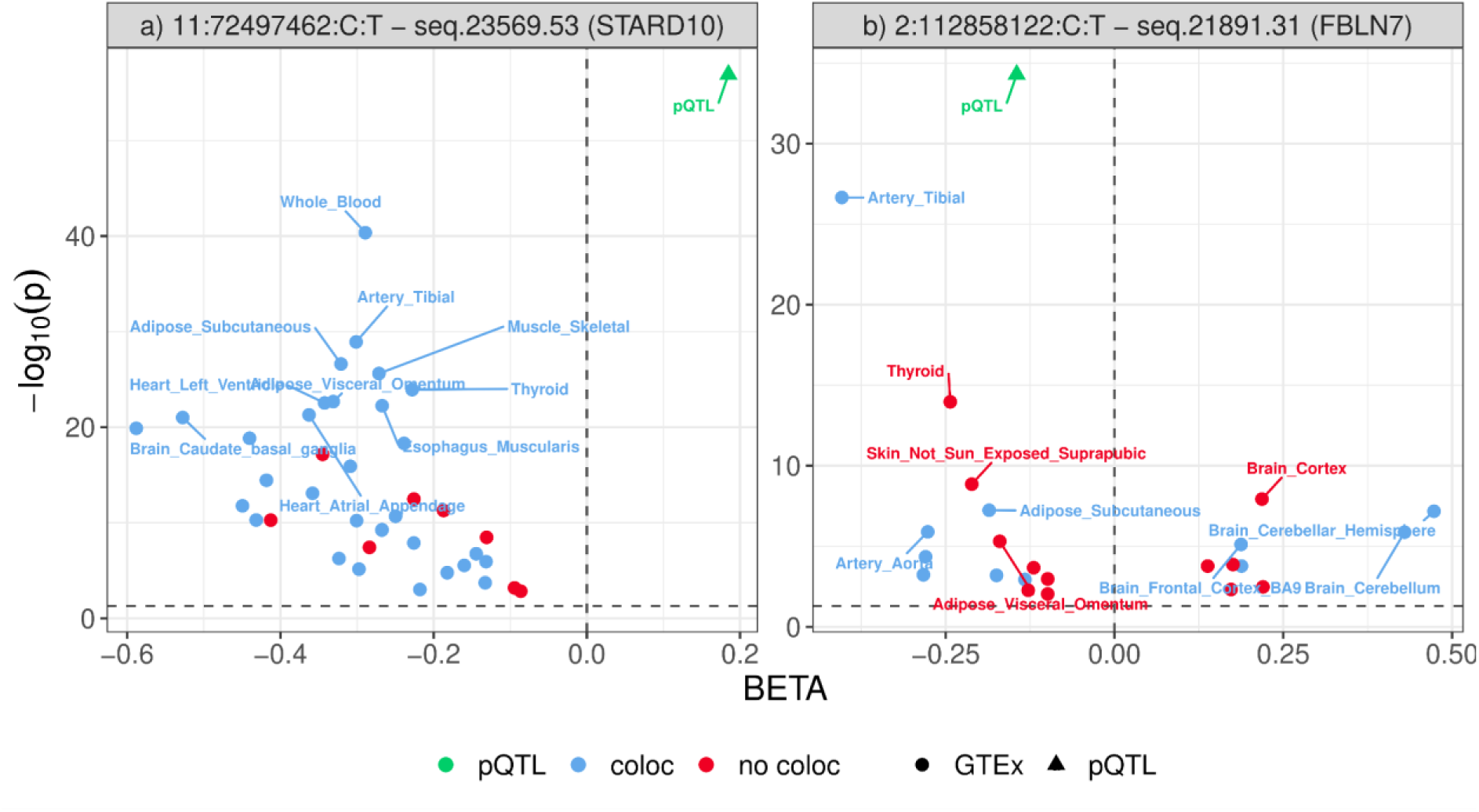
Examples of discordant pQTLs and eQTLs. For the respective index pQTL and all GTex tissue eQTLs that were significant at a 5% FDR threshold, the -log10 P-value is displayed against their beta (effect). The green triangle represents the index pQTL, blue points indicate significant eQTLs where the respective regional pQTL association significantly colocalizes (PP.H4 > 0.8) with the gene expression data of the encoding gene in that tissue, red points indicate significant eQTLs where the respective regional pQTL association does not colocalize with the gene expression data of the encoding gene in that tissue. If the effect allele tested in GTEx corresponded to the non-effect allele in this meta-analysis, the sign of the GTEx effect was inverted. a) pQTL 11:72497462:C:T - seq.23569.53, encoding gene *STARD10*, MAF=0.34, GTEx tissues that have an eQTL of - log10(p) > 20 are labelled; b) pQTL 2:112858122:C:T - seq.21891.31, encoding gene *FBLN7*, MAF=0.41, GTEx tissues that have an eQTL of -log10(p) > 5 are labelled.

**Figure S11.**
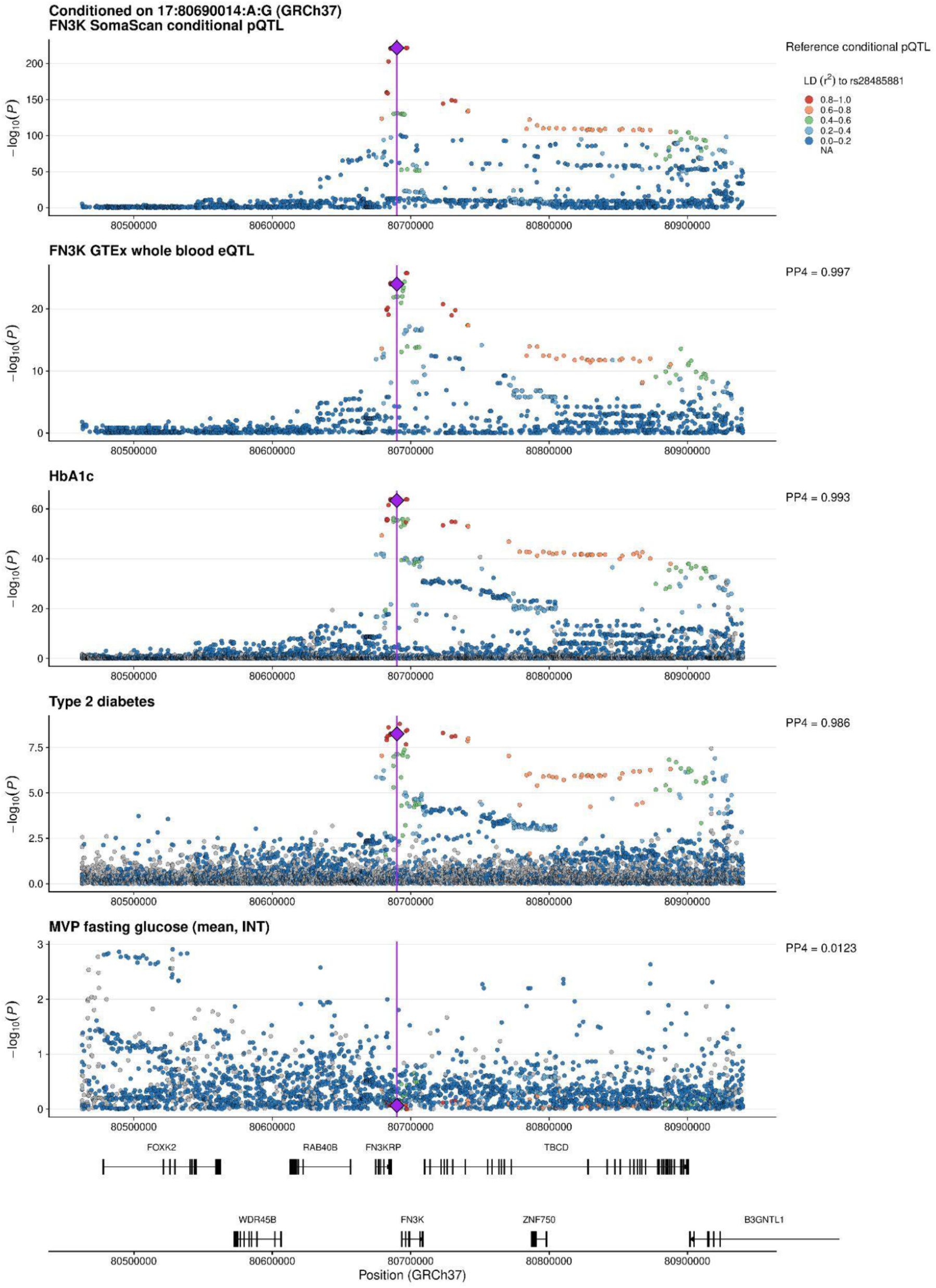
Regional association plots across the FN3K locus. From top to bottom: conditional association for circulating FN3K plasma protein levels (INTERVAL–CHRIS meta-analysis; N = 13,445); FN3K whole-blood expression (GTEx v8 whole blood; N = 1,116); HbA1c (UK Biobank + MVP meta-analysis; UK Biobank N = 400,837; MVP N = 429,670); type 2 diabetes (UK Biobank + MVP + FinnGen meta-analysis; UK Biobank: 22,768 cases/418,949 participants; MVP: 156,658cases/463,989 participants; FinnGen: 65,085 cases/400,197 participants); and fasting glucose (MVP EUR-HARE; inverse normal-transformed mean fasting glucose; N =16,316). All association statistics are displayed across the same genomic region (GRCh37). The primary conditional cis-pQTL variant, rs28485881 (GRCh37: 17:80690014:A:G), is indicated by the purple diamond and vertical line and was used as the reference variant for linkage disequilibrium (LD) estimation. Point colours represent INTERVAL LD (r²) with rs28485881. Posterior probabilities of colocalization supporting a shared causal variant (PP.H4) are shown to the right of each panel. The bottom panel shows GENCODE v19 (GRCh37) protein-coding gene models, with one representative transcript displayed per gene.

**Figure S12.**
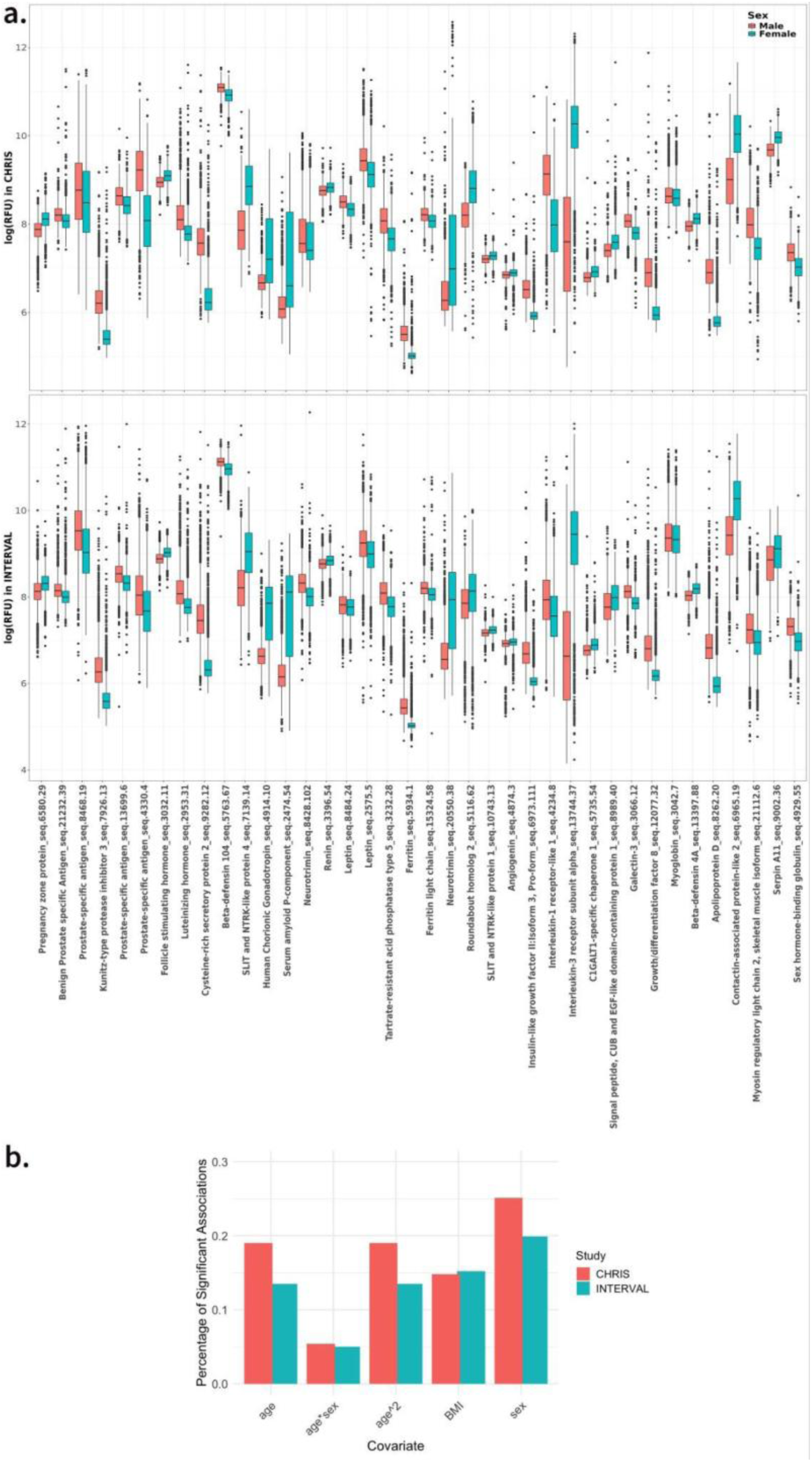
Non genetic associations. (**a**) Distribution of unadjusted protein levels by sex (Relative Fluorescence Units - log scale) of 38 aptamers targeting sex-specific proteins (with corresponding protein name) in INTERVAL and CHRIS. (**b**) Each aptamer was tested in the INTERVAL cohort for association with sex, age, age², BMI, and the age–sex interaction using linear regression. Bars show the proportion of proteins significantly associated with each covariate after multiple testing correction.

**Figure S13.**
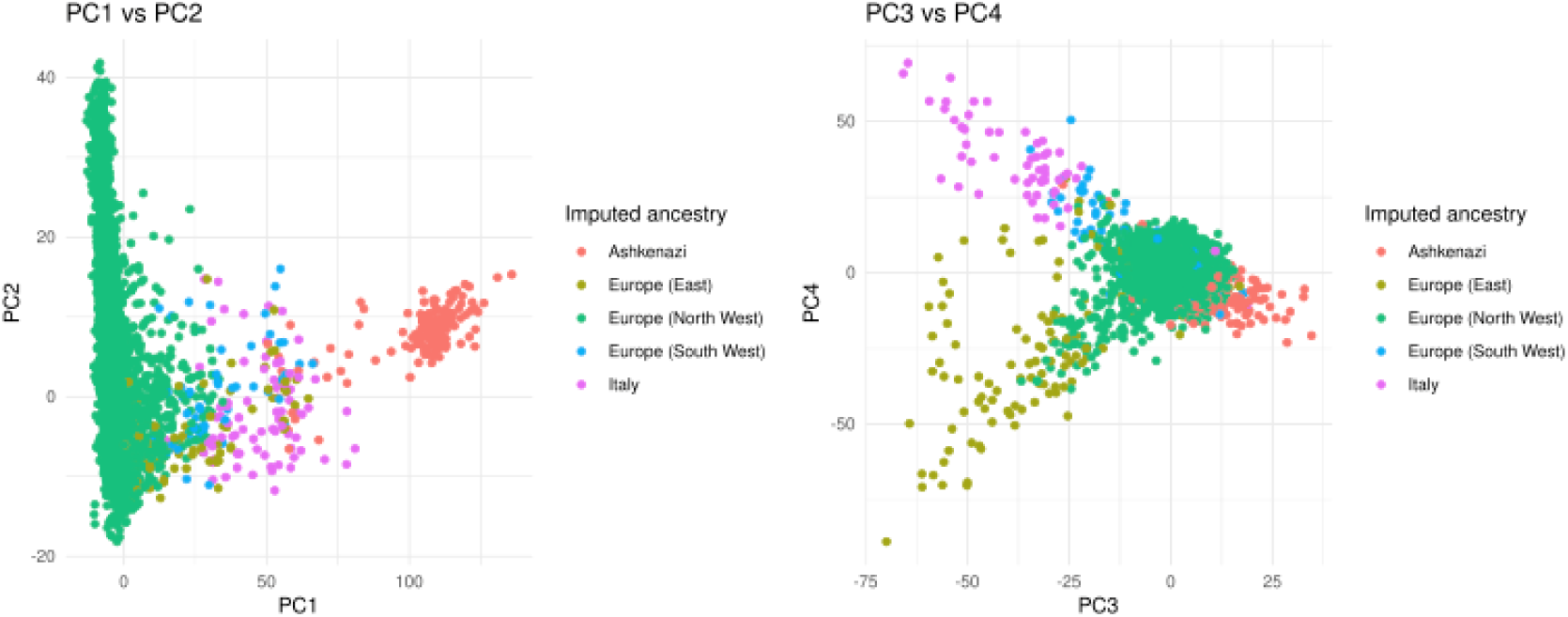
PCA-based ancestry inference. Principal components were computed in the INTERVAL cohort and colored by ancestry imputed from allele frequencies, using projection onto UK Biobank reference groups as described in Privé (2022).

## Supplementary Tables

**Supplementary Table 1: List of aptamers covered with annotation, LOD and CV.** Legend: Lists N=7,289 aptamers considered in each study, before LOD filtering, along with their associated annotations and assay details. For each aptamer, we provide the limit of detection (LOD), coefficient of variation (CV), transcription start site (TSS), UniProt identifier, and additional information on the assay used. These annotations facilitate quality assessment, reproducibility, and interpretation of the aptamer-based measurements.

**Supplementary Table 2: Regional associations annotated.** Legend: Loci (N= 7,870) identified as significant regional pQTL associations (aptamer-locus pairs). For each locus, key information including the most significant (lead) variant, chromosomal coordinates, effect sizes, and significance levels are provided. The table also includes functional annotations such as nearby genes, regulatory features, and relevant biological insights to aid in interpreting the genetic associations. These annotations support the identification of candidate regions for further investigation.

**Supplementary Table 3: Conditional SNPs associations annotated.** Legend: List of 12,095 conditionally independent variants at each of the 7,870 regional associations (SeqID-LOCUS) annotated by Ensembl Variant Effect Predictor (VEP). Conditional model coefficients are represented with “-” if the SNP does not have any independent variant in the COJO-conditional analysis. The table also shows 99% credible set variants (posterior inclusion probability, PIP > 0.99) for each of the 12,095 independent variants, resulted from coloc fine-mapping analysis (see Methods), and indicates any protein-coding genes affected by moderate- or high-impact conditionally independent variants or their proxies at the locus.

**Supplementary Table 4: Determinants of *cis*-pQTL discovery and *cis*-regulatory complexity. Logistic regression models of *cis*-pQTL discovery and *cis*-regulatory complexity.**

Legend: Logistic regression results testing biological and technical determinants of *cis*-pQTL detection across all quality-controlled SomaScan aptamers (*cis* discovery models) and determinants of *cis*-regulatory complexity among targets with at least one *cis*-pQTL (*cis* complexity model). Models report log-odds.

**Supplementary Table 5: Summary of the 22 identified genomic hotspots.** Legend: For each hotspot, the table reports chromosomal position, genomic window (GRCh37), counts of *cis* and *trans* signals, number of colocalized connected communities, protein-coding genes with *cis* signals, overlapping genes, and the most frequent SNP in colocalizations.

**Supplementary Table 6:List of 2,003 phenotypes used as outcomes in the Mendelian Randomization (MR) analyses.** Legend: Summarizes the 2,003 phenotype definitions used as outcomes in the Mendelian Randomization (MR) analyses and reports case–control counts across multiple biobanks and genomic resources. Each row corresponds to a harmonized phenotype and includes curated descriptions from the Million Veteran Program (MVP), UK Biobank (UKBB), and FinnGen, together with the associated UKBB phenocode and cohort-specific sample-size information. Metadata on data sources and the standardized phenotype classification assigned to each trait are also provided.

**Supplementary Table 7: List of significant MR traits with identified opportunities for drug retargeting.** Legend: A total of 2,003 traits were initially evaluated using Mendelian Randomization. After applying the stringent genome-wide significance threshold (P-value < 1.43 × 10⁻⁸) to account for multiple testing, 643 traits demonstrated significant associations with the outcome. Among these, 751 corresponded to rediscoveries of known drug indications, while 367 represented potential repurposing opportunities for alternative indications.

**Supplementary Table 8: Sex-specific associations of 39 raw proteins levels with sex in INTERVAL and CHRIS.** Legend: The list of proteins and corresponding aptamers expected to be associated with biological sex was provided by SomaLogic.

**Supplementary Table 9: Non-genetic associations.** Legend: Adjusted R² values from the regression model explaining the proteomics principal components (PCs) based on various sets of factors. The final model used to adjust the protein measurements is highlighted in blue.

