## supplementary methods for "Beyond the classical plasma secretome: genetic architecture and disease associations of the expanded human plasma proteome in 13,445 Europeans"

**Title**

—------------------------------------------------------------------------------------------------------------------

### Supplementary Methods

#### SM1. Study cohorts

##### SM1.1 INTERVAL cohort participants

The INTERVAL study[1] consists of nearly 50,000 whole blood donors recruited between 2012 and 2014 into an open randomised pragmatic trial of varying blood donation intervals. Recruitment took place at 25 centres of England's National Health Service Blood and Transplant (NHSBT). All participants provided informed consent prior to joining the study and the National Research Ethics Service approved this study (11/EE/0538). All participants, who were aged 18-80 at recruitment, were in generally good health as people with major diseases (myocardial infarction[MI], stroke, cancer, HIV, and hepatitis B or C) or who had recent illness or infection would not have been eligible to donate blood. Non-fasted blood samples were collected at the beginning of a regular blood donation session, providing whole blood, serum and plasma for bioassays. Participants were then asked to fill in online questionnaires regarding their lifestyle, demographic characteristics, anthropometry (including height and weight) and diet. 9,769 participants were selected using a case-cohort design for assay using the SomaScan platform v4.1. Suspected incident cases of several diseases (diabetes, end stage renal disease, MI, lung disease, heart failure, stroke, blood cancers, primary and secondary malignancies) were selected and controls were randomly selected after case identification.

##### SM1.2 CHRIS cohort participants

The Cooperative Health Research in South Tyrol (CHRIS)[2,3] study is a population-based study conducted in the Vinschgau/Venosta district in South Tyrol (Italy) to investigate the genetic and molecular basis of age-related conditions and their interaction with life-style and environment in the general population. From 2011 to 2018, 13,393 adult residents (≥18 years) participated in the study. Following overnight fasting, participants underwent blood drawing, urine collection, anthropometric and blood pressure measurements, as well clinical measurements and questionnaire-based interviews to screen for cardiovascular, renal, endocrine, metabolic, neurological, behavioural and cognitive system conditions, and to assess lifestyle exposures including diet, physical activity, and life-course smoking. 4,229 participants were selected for assay using the SomaScan platform v4.1: samples were selected to maximise overlap with other available resources, specifically with targeted metabolomics collection, proteomics SWATH collection and the GA2LEN food frequency questionnaire. The study was approved by the Ethics Committee of the Healthcare System of the Autonomous Province of Bolzano/Bozen - South Tyrol (21/2011). All CHRIS study participants provided informed dynamic consent (PMID: 36064788). This specific data use was part of the general consent for the study, but participants were provided with specific information on the research through an individual communication, providing the opportunity for opting out of this specific data use.

### SM2. Protein assays and quality control
#### SM2.1 Protein assays and quality control in INTERVAL

We used the 7k version (v4.1) of the multiplexed aptamer-based approach SomaScan assay to quantify plasma levels of proteins in the subset of 9,769 participants from the INTERVAL substudy. Aliquots of 55 µl of plasma were sent to SomaLogic Inc. (Boulder, Colorado, US) in 2 pre-randomized batches for "SomaScan" protein measurement. Each batch was assayed independently using a previously described protocol[4]. Briefly, modified fluorescent DNA aptamers are used to bind to specific proteins and are then hybridized to a DNA chip for quantification. Protein abundances were quantified using SomaScan's aptamer-based assay, where multiple aptamers can sometimes target the same protein.

Harmonization for within-run and between-run technical variabilities using hybridization controls on the microarray and calibrator samples was performed by SomaLogic within each batch. 57 samples in batch 1 and 55 samples in batch 2 did not pass internal acceptance criteria because of extreme calibration scale factors or technical problems recorded during the assay, leaving measurements of 7,289 aptamers for 9,657 participants (4,933 in batch 1 and 4,724 in batch 2) **(Supplementary Table 1)**.

Quality control was performed separately within each batch. Coefficients of variation (CV) were computed for each aptamer on the 346 (176 in batch 1 and 170 in batch 2) quality control samples that passed SomaLogic's internal acceptance criteria. 108 aptamers in batch 1 and 121 aptamers in batch 2 with a high CV (≥20%) were flagged in our dataset but still included in our analyses. The limit of detection (LOD) was computed per aptamer in each batch using measurements from buffer samples. LOD was defined per aptamer per batch as:

LODaptamer = median(Buffer measurementsaptamer)+5xMAD(Buffer measurementsaptamer)[4]

where MAD refers to Median Absolute Deviation. 1,826, 1,264 and 2,005 human aptamers had at least one value below LOD in batch 1, batch 2 or either batch, respectively. We excluded 144 aptamers with more than 50% of values below LOD in at least one batch, leaving measurements of 7,145 aptamers in 9,657 participants, from which we removed 2 participants who had withdrawn from the INTERVAL study **(Supplementary Table 1)**. For non-genetic analyses, we used SomaLogic ANML-processed data, log-transformed and scaled.

#### SM2.2 Protein assays and quality control in CHRIS

Protein measurements were performed following the exact same assay and protocol as for INTERVAL by SomaLogic on a single pre-randomized batch of 4,229 samples. Samples were selected to maximize overlap with other datasets in CHRIS (targeted metabolomics, scanning SWATH proteomics, and GA2LEN food frequency questionnaire). Measurements were obtained for 7,289 aptamers corresponding to 6,381 proteins for 4,204 participants after removal of the samples not passing SomaLogic's internal acceptance criteria **(Supplementary Table 1)**.

CV and LOD were computed as in INTERVAL, with 167 aptamers having a CV > 20% (no aptamer removal based on CV). Aptamers with more than 80% of samples below LOD were discarded, retaining measurements of 7,261 aptamers for 4,204 participants **(Supplementary Table 1)**.

##### SM2.3 Comparison of proteomics measurements across batches

Reproducibility of measurements, as assessed by coefficients of variation (CVs) in technical replicates, was high with median coefficient of variation (CV) in the 3 batches of 4.1%; 4.4%; 4.8%. We found high between-batch correlation of CVs and strong correlation with a previous study[4] (**Supplementary Figure 4**). Uncorrelated outliers were largely driven by the dilution factor used within the assay (**Supplementary Figure 4**). Per-aptamer limits of detection (LOD) estimated from buffer samples were also highly correlated ($r^2$>0.75) across batches (**Supplementary Figure 5**. The number of aptamers for which more than 20% of samples were below the LOD ranged from 158 (CHRIS) to 251 (INTERVAL batch 1). We replicated associations with biological sex in both INTERVAL and CHRIS of raw levels 38 aptamers previously known to be sex-associated, including pregnancy zone protein (PZP) and prostate-specific antigen (PSA). All 38 aptamers (one was excluded from testing as it failed QC) were significantly associated with sex in both INTERVAL (P-value<0.0026, adjusted on batch) and CHRIS (P-value<0.0008) after Bonferroni correction for multiple testing (**Supplementary Figure 12a, Supplementary Table 8**).

We also established the contribution of other biological factors (age, $age^2$, body-mass index and the interaction between sex and age) to individual aptamers raw levels in both INTERVAL and CHRIS. Through a regression model we assessed the significance of each of the above factors to predict aptamers' expression, accounting for multiple testing. We report the percentage of aptamers significantly associated with each factor **(Supplementary Figure 12b)** and observe that all factors are significantly associated with a large percentage of individual aptamers coherently between the two cohorts.

##### SM2.4 Associations of proteomics data with technical and biological factors in INTERVAL

In INTERVAL, we assessed the impact of a broad range of biological (age, body-mass index, C-reactive protein, smoking status, the first ten genetic principal components, and case labels) and technical (assay plate, batch, time from blood draw to sample freezing, time and season of the blood sampling) factors on the first five principal components (PCs) computed on INTERVAL's log-transformed standardized aptamer levels, in order to understand what drove variability in proteins the most (**Supplementary Table 9**). The first protein PCs were predominantly driven by technical variability, particularly plate ($R^2$ = 31.4%, 29.0%, 17.2% for PC1–PC3), processing delay ($R^2 \approx$ 9.3% for PC1–PC2), and batch ($R^2$ = 3%, 5.4% for PC1 and PC3) with smaller effects from time of day ($R^2 \approx$ 2–3%). In contrast, biological factors explained negligible variance (< 1%). Multivariable models confirmed that including technical factors was essential to adjust for systematic variation. Conversely, models with only biological and sample collection-related covariates explained limited variance (adjusted $R^2 \approx$ 0.12–0.14 for PC1-PC2).

#### SM3. Genetic data processing and quality control

##### SM3.1 INTERVAL cohort

The genotyping and imputation protocol for the INTERVAL samples has been described previously in detail[5]. All participants were genotyped on the ThermoFisher UK Biobank genotyping array. We performed additional quality control on both genotyped and imputed datasets specifically on the subset of 9,327 participants with both proteomic and genomic data. We excluded samples identified as outliers in terms of population ancestry and samples with extreme heterozygosity.[6] We used only autosomal chromosomes. Multi-allelic variants were re-coded as pseudo bi-allelic.

We had data for 43,059 samples and 655,045 variants in our raw genotype file. We first kept only the 9,327 samples for which we also have SomaScan proteomics data. We then identified the outliers in term of heterozygosity: we used PLINK 2.0 to restrict to the autosomal chromosomes, and filter out for low call rate (samples and variants, threshold 0.1), for not passing Hardy Weinberg equilibrium test (threshold $10^{-15}$) and for minimum allele frequency lower than 0.01. We then computed heterozygosity using PLINK and kept only samples with F coefficient within 3 standard deviations of the population mean[6], leading to excluding 72 samples.

Genetic principal components were computed from the genotype file after excluding variants associated with population structure[7]. Variants associated with population structure were identified by projecting the genotype matrix onto its first five principal components, following the procedure described in The UK Biobank resource with deep phenotyping and genomic data[8], and detected using the bed_pcadapt function from the *bigsnpr* package[9] to identify and remove genetic markers involved in biological adaptation, ending up with 495,110 variants.

We used PLINK to compute relatedness from this set of variants, using a Kinship threshold of $2\times10^{-4.5}$ and then excluded the 59 individuals found as related to restrict to relatedness lower than 3rd degree (strict exclusion of all pairs of related samples), as determined using *KING*[10] without LD pruning. Principal components were then re-projected onto the full sample set, including the related individuals.

We performed PCA on the set of 9196 unrelated samples for the same 495,110 variants as before, using the R *bigsnpr* package[9] which allows removal of long-range LD regions and automatic LD clumping (we used the default parameters) and then reprojected our PCs on our whole population including the related samples.

To infer ancestry, we also projected our data onto the PCA space of many known population groups defined from UK Biobank.[11] Looking at the first components, we decided to exclude 2 outliers which were inferred to be Middle East ancestry, whereas the majority of our population seems to be European or Ashkenazi **(Supplementary Figure 12)**, ending up with 9,253 samples. Removing 2 samples which dropped out from the cohort, we obtained our final list of 9,251 individuals to be kept in the genotyped data. We restricted the initial raw file to these samples, and applied again the autosomal chromosomes, low call-rate, missingness and Hardy-Weinberg equilibrium filters described above, ending up with 594,319 variants. We computed the genetic principal components used later in the analysis on this updated file following the exact same method as described above.

Methods for imputation and additional QC steps performed prior to imputation have been described extensively before.[5] Briefly, strong variants filtering was performed to prepare a high quality imputation scaffold. The dataset was phased using SHAPEIT3 and imputed using the

1000 Genomes Phase 3-UK10K imputation panel[12] on the Sanger Imputation Server (https://imputation.sanger.ac.uk), using the PBWT algorithm.[13]

We had data for 43,059 samples and 87,696,888 variants. We restricted our analysis to the 9,251 samples from the INTERVAL study identified from genotype data files. We first used PLINK 2.0 to recode the variants ID to chromosome:position:allele1:allele2, especially to handle multiallelic variants (now coded as pseudo-biallelic); and to filter out sex chromosomes, samples with low call-rate (threshold: 0.1), variants not passing Hardy Weinberg Equilibrium test (threshold for P-value: $10^{-15}$) and variants with low allele count (threshold: 10). We computed an imputation quality metric (INFOscore) through QCtools and kept only variants with INFOscore higher than 0.7, obtaining a final set of 13,749,088 variants. We converted these harmonized PGEN files to hard-called binary PLINK 1.9 (bed/bim/fam) format and later employed them to compute the reference LD panel for regional association plots and for COJO-conditional analyses.

##### SM3.2 CHRIS cohort

Genotyping was performed using the Illumina HumanOmniExpressExome and Illumina Omni2.5Exome arrays. Illumina GenomeStudio v2010.3 with default settings was used to call genotypes on GRCh37. Variants with GenTrain score < 0.6, cluster separation score < 0.4, or call rate < 80% were considered technical failures and discarded. Only variants present on both arrays were forwarded to our standard quality control pipeline. Samples with a call rate < 98%, monomorphic variants or variants with Hardy-Weinberg equilibrium P-value < $10^{-6}$ were removed. Genotypes were imputed using the Michigan Imputation server with the Haplotype Reference Consortium (HRC) reference panel on the genome in build GRCh37. SNPs with low imputation quality scores (INFO<0.7) and MAC<10 were excluded from the analysis. Multi-allelic variants were split into multiple lines, left-aligned and normalized using `bcftools norm`, large insertions and deletions were removed from the GWAS analysis.

#### SM4. Genetic association analyses

##### SM4.1 GWAS analyses

Protein measurements were provided by SomaLogic as part of the “Adaptive Normalization by Maximum Likelihood” (ANML)–processed dataset, which applies quality control and median normalization across QC and clinical samples to correct for technical variation, including plate-to-plate variability. We then performed quality controls independently in all 3 batches (2 batches for INTERVAL, 1 batch for CHRIS, see **SM2**). Protein abundances were then inverse-rank normalized prior to genetic association testing.

GWAS analyses were performed independently in our 2 cohorts using REGENIE (v3.3) implemented in Nextflow (v. 22.10.1). Through a 2-step procedure, REGENIE[14] uses polygenic effect estimates to control for population and relatedness. In brief, the first step consists of a whole-genome regression model using a Leave-One-Chromosome-Out (LOCO) scheme to capture the proportion of phenotype variance explained by genome-wide polygenic effects; the second step performs single SNP association tests using linear regression for quantitative traits, conditional on the prediction from the first step model. For the first step, we used the genotyped (i.e. non-imputed) cleaned dataset as a set of high-confidence SNPs. Based on analysis of technical factors associated to aptamers’ variability (**SM2.3 and SM2.4**), the association models in INTERVAL included the following covariates measured on 9,251 samples and 7,145 proteins: batch, age, sex, time between blood draw and processing, season and the first 10 genetic principal components (PCs) (**Supplementary Figure 13**). We decided to include batch instead of plate since the former grouped the latter, and we aimed at the simplest model. In CHRIS, we used the same covariates (except batch) measured on 4,194 samples and 7,261 proteins. In both cohorts, linear residuals were computed and then were used as input for REGENIE. To assess overdispersion in each GWAS, we computed the genomic inflation factor using the median of the resulting chi-squared test statistics divided by the expected median of the chi-squared distribution.

#### SM5. Harmonization and meta-analysis

We used METAL[15] to perform an inverse variance weighted meta-analysis, with estimates of heterogeneity, using GWAS results of the 7,144 aptamers retained after QC in both cohorts. Allele matching was automatically performed by METAL. INTERVAL was the reference study for sign. Only variants common to the 2 studies were kept in the meta-analysis summary statistics for downstream analyses, which resulted in 8,613,862 variants (INFO > 0.7). Between-study heterogeneity was quantified using Cochran's Q statistic and the $I^2$ metric.

#### SM6. Regional associations

For most of the downstream analyses, we used the conventional genome-wide statistical significance threshold (P-value < $5\times10^{-8}$) adjusted for multiple testing to account for the large number of proteins. We computed an effective number of tests for Bonferroni adjustment using the number of PCs needed to account for 95% of the variability of the protein dataset in each cohort. We selected the value from INTERVAL (3,978) as it was the most conservative. We hereafter used "our Bonferroni-corrected significance threshold" or *p1* to refer to the threshold $1.26\text{x}10^{-11}$ (i.e. $5\times10^{-8}$/3,978).

To identify significant genomic regions for each aptamer in our GWAS meta-analysis, we first selected all variants with a P-value < $10^{-6}$. These SNPs were grouped in a genomic region ('locus') if they were located on the same chromosome and within 3 million base pairs of each other.[16] A locus was considered significant if it contained at least one SNP with a P-value < $1.26\text{x}10^{-11}$. For each aptamer, we counted the number of significant loci, with the total across all aptamers being 9,460 significant locus–aptamer associations.

We excluded from downstream analyses two loci we considered to be problematic for fine-mapping: the *HLA* region (chromosome 6, positions 28,477,797 to 33,448,354 in build GRCh37), due to its complex pattern of linkage disequilibrium (LD), and an extended region around the *NLRP12* gene (chromosome 19, positions 54,300,000 to 54,360,000 in build GRCh37), which has been previously identified as a region containing signals specific to the INTERVAL study[17]. After excluding these loci, 7,870 significant locus-aptamer associations remained for downstream analyses.

#### SM7. Protein-to-gene mapping and identification of *cis/trans* signals

To be able to define *cis* and *trans* association signals, we mapped the aptamers to the transcription start site (TSS) of the corresponding target protein(s). First, we retrieved a table containing 7,596 aptamers and their corresponding Entrez Gene ID and UniProt IDs from SomaLogic menu download (version 4.1, 7k panel, retrieved November 26, 2024, from https://menu.somalogic.com/ — access login-restricted). After removing non-human and non-protein aptamers, 7,289 aptamers were left for TSS extraction. Of the 7,289 aptamers, 82 mapped to more than one UniProt ID, and 88 mapped to more than one Entrez Gene ID. Splitting these entries into unique aptamer - UniProt - Entrez Gene ID triplets resulted in 7,397 combinations for TSS annotation. Using the NCBI feature table for GRCh37 (https://ftp.ncbi.nlm.nih.gov/genomes/all/GCF/000/001/405/GCF_000001405.25_GRCh37.p13/GCF_000001405.25_GRCh37.p13_feature_table.txt.gz), genes in the SomaLogic table were matched to NCBI via the Entrez Gene ID using feature='gene', class='protein_coding' and assembly_unit='Primary Assembly' preferentially and only to non protein-coding genes if no protein coding gene existed. To determine the TSS, we took the 'start' position as the TSS if the strand was positive and the 'end' position if the strand was negative.

A mapping of UniProt ID to Entrez Gene ID was obtained from the UniProt database (retrieved November 26th, 2024) to validate consistency of UniProt to Entrez Gene ID provided by SomaLogic by matching according to UniProt. For aptamers with multiple UniProt - Entrez Gene ID pairs, where exactly one pair matched according to the UniProt database, we removed the TSS annotation for the non-matching pairs. This procedure removed one annotation each for two aptamers and three annotations for one aptamer. For 7,194 aptamers, exactly one TSS was assigned, for 73 aptamers, two TSS were assigned and for the remaining 12 aptamers more than two TSS were assigned. For 10 aptamers, no TSS could be automatically assigned. These cases were manually curated using information from GeneCards (https://www.genecards.org/), taking into account gene proximity, annotation consistency, and haplotype variation. When multiple nearby genes were present, the most representative locus was selected based on genomic context and strand orientation. In cases where gene identifiers were discontinued or unavailable in the reference genome, alternative names or equivalent loci were identified through GeneCards to ensure consistent TSS assignment. For the unique remaining aptamer without an assignable TSS, the chromosome value was set to 0 and the TSS position to –999,999 to avoid missing (NA) entries in downstream analyses.

The Ensembl Gene IDs were matched using the Ensembl biomaRt in R. Matching was performed via the Entrez Gene ID where possible and by UniProt ID where no match was found by Entrez Gene ID. For downstream analyses, Entrez Gene IDs were further mapped to HGNC gene symbols using the mapIds function from the *AnnotationDbi* R package (version 1.68.0), in conjunction with the *org.Hs.eg.db* annotation package for *Homo sapiens* (version 3.20.0). Entrez Gene IDs were first converted to character format to ensure compatibility with the mapping function. Gene symbols were retrieved using the SYMBOL column with ENTREZID as the key type, and in cases where multiple mappings were possible, only the first match was retained. After removal of non-human aptamers, resolution of inconsistent mappings, and manual curation of TSS assignments, the final annotation dataset (**Supplementary Table 1**) contained 7,391 aptamer–protein-genes combinations corresponding to 7289 unique aptamers, 6381 proteins, 6379 genes.

For each aptamer, we then defined the corresponding *cis* region(s) as a window of +/-500kb around the TSS(s). A regional association was defined as *cis* if the corresponding locus overlapped any *cis* region of the associated protein, and as a *trans* association otherwise.

#### SM8. Overlap of aptamers and proteins with previous the SomaScan assay platform

To determine whether each SomaScan aptamer was newly assessed in the 7k version (v4.1) of the assay, we used the SomaScan.db R package (v4.0 panel), since previous versions are no longer available from the SomaLogic menu. Aptamers were matched using SeqId, and those already present in v4.0 were flagged as previously assayed. Although minor differences exist between the v4.0 mappings from SomaScan.db and the 7k mapping obtained from the SomaLogic menu (72 mismatched UniProt entries and 58 mismatched Entrez Gene IDs), these were minimal and likely reflect annotation updates. For consistency, the menu-derived mapping was retained for all downstream analyses.

Based on the 7k aptamer-to-UniProt mapping, we flagged each aptamer (by SeqId) and its corresponding protein (by UniProt ID) for prior inclusion in SomaScan v4.0.

Proteins were classified as previously available or newly added based on their complete representation in this earlier SomaScan assay version. Specifically, we first annotated each aptamer according to whether it was included in the SomaScan v4.0 (≈5k) assay. Proteins (UniProt IDs) were then classified in the following way: a protein was defined as new in the 7k panel only if none of its corresponding aptamers were present in the prior SomaScan 5k assay. Conversely, a protein was considered previously available if at least one aptamer targeting that protein was already included in the 5k panel (**Supplementary Figure 13**). This strict protein-level definition ensures that "newly added" proteins represent genuine expansion of the detectable plasma proteome rather than alternative assay reagents targeting previously measured proteins.

#### SM9. Protein biological annotations

##### SM9.1 Annotation of plasma proteins with secretion pathway

Plasma proteins were annotated according to their likely mechanism of entering the bloodstream[18]: (1) actively secreted to blood, (2) cleaved from the cell membrane, and (3) released from intracellular compartments (e.g., cytoplasm or nucleus) due to cell damage or leakage (**Supplementary Table 1**) . Proteins that are actively secreted to blood were annotated using the manually curated list of the human secretome by Uhlen, M et al. 2019[19] which identified 1709 proteins secreted to blood or local tissues. The proteins cleaved from the cell membrane were annotated using the "cell membrane" keyword from the subcellular location information in the UniProt database.[20] The remaining proteins were considered to enter the bloodstream due to cell leakage, coming from the intracellular space. Proteins with isoforms linked to multiple secretion mechanisms were annotated in order of priority: (1) actively secreted to blood, (2) cleaved from the cell membrane, and (3) cell leakage; thus insuring that the mechanism most likely to explain measurement in blood was prioritized In figures and counts, for proteins with multiple aptamers, the most frequent category (mode) was used. Percentages were calculated after excluding proteins with missing annotations.

**SM9.2 RNA tissue annotations (Human Protein Atlas)**

We annotated each protein (UniProt ID) in our dataset with tissue-level RNA expression categories using the Human Protein Atlas (v24) (**Supplementary Table 1**) . Expression data in HPA are defined at the gene level (HGNC gene symbols), with UniProt accession numbers provided as secondary mappings. The RNA expression categories are derived from a consensus integration of HPA's own RNA-seq data with GTEx and FANTOM5. We exported the HPA *tissue category* table (fields: *RNA tissue specificity*, *RNA tissue distribution*, *RNA tissue specific nTPM*) and the *consensus RNA expression* table (rna_tissue_consensus.tsv, nTPM values across 50 tissues). We joined HPA annotations to our dataset by gene symbol, and aggregated to the UniProt level using the mode (most common value across aptamers mapping to the same protein). Entries labelled 'Not detected', 'NA', empty, or 'N/A' were treated as missing and excluded prior to computing percentages.

For each protein, we extracted the following fields:

1. **RNA tissue specificity** — tissue enriched, group enriched, tissue enhanced, low specificity, or not detected. According to the Human Protein Atlas, "tissue enriched" denotes at least four-fold higher mRNA expression in one tissue compared with any other tissue; "group enriched" denotes at least four-fold higher average mRNA expression in a group of 2–5 tissues compared with any other tissue; and "tissue enhanced" denotes at least four-fold higher mRNA expression in one tissue compared with the average across all other tissues. "Low tissue specificity" indicates expression (nTPM ≥ 1) in at least one tissue without evidence of elevation in any tissue, and "not detected" indicates nTPM < 1 in all tissues.

2. **RNA tissue distribution** — detected in all, many, some, single, or not detected.According to the Human Protein Atlas, "detected in all" indicates detection in all tissues; "detected in many" indicates detection in at least one third but not all tissues; "detected in some" indicates detection in more than one but fewer than one third of tissues; and "detected in single" indicates detection in a single tissue.

3. **RNA tissue specific NX** — the HPA-provided *"RNA tissue specific nTPM"* field, which reports quantitative expression values for enriched/enhanced tissues, formatted as "tissue: value; tissue: value".

4. **RNA tissues detected (nTPM ≥ 1)** — derived from the consensus RNA expression table as the list of tissues where the gene was expressed above the HPA detection threshold.

Among the UniProt accessions analysed, the vast majority showed concordant mapping to HPA gene symbols.

##### SM9.3 Blood concentrations (Human Protein Atlas)

Expected absolute protein concentrations in human plasma (pg/L) were retrieved from the HPA blood protein dataset (mass spectrometry-based). Ensembl gene IDs were mapped to UniProt accessions using HPA's proteinatlas.tsv file. Concentration values were log10-transformed for statistical comparison between groups. We then compared expected plasma concentrations between proteins already present in earlier SomaScan versions and those newly added in the 7k version. Entries where no blood concentration was available were coded as NA (**Supplementary Table 1**) .

#### SM10. Determinants of pQTL discovery

Logistic regression models were fitted at the aptamer level using the glm() function in R with a binomial link function. The primary outcome was the presence of at least one *cis*-pQTL per aptamer. Predictor variables included assay detectability (percentage below LOD in distinct batches), assay precision (mean CV), protein secretion pathway, expected plasma protein concentration ($log_{10}$-transformed), and aptamer panel novelty. Both univariate and multivariable models were fitted; the multivariable model jointly included assay novelty, assay detectability, secretion pathway, and plasma concentration. All models were restricted to quality-controlled aptamers with complete data for the covariates included in each model (complete-case analysis). No imputation was performed, and results are reported as odds ratios with corresponding Wald P-values.

#### SM11. Identification of genetic hotspots of regional associations

We identified genomic regions with a high density of associated proteins, which we termed "hotspots". To delineate these hotspots, chromosomes were systematically partitioned into contiguous windows of 500 Kb. Within each window, we quantified the number of overlapping signals by evaluating whether the genomic loci associated with each signal intersected the window boundaries. Windows with more than 50 signals were classified as hotspots. Subsequently, contiguous windows meeting hotspot criteria were merged to define broader hotspot regions, facilitating downstream biological analyses.

#### SM12. Determinants of heterogeneous signals.

Heterogeneous signals were defined as signals with I2 higher than 90% for the most significant SNP. 2236 (36%) regional associations were thus identified as heterogeneous. 78% (2236) signals were located within a genetic hotspot.

Outside hotspots, 608 out of 3447 signals (18%). were heterogeneous. More than one third of them (237; 39%) were strong (meta-analysis P-value > $10^{-40}$ ) *cis* signals, significant in both studies and with consistent effect directions, while 60 of them showed different allele frequencies (minimum allele frequency difference > 5%) in the 2 studies. For the remaining

335 ones, we compared study-specific P-values, allele frequencies, and CV distributions to all 3447 signals outside hotspots. Unexplained heterogeneous signals were more likely to be not significant in one of the 2 studies (244 signals with at least one of the study-specific 1.26 × $10^{-11}$, $Chi^2$ P-value = 9 x $10^{-7}$), and more likely to have MAF<0.05% ($Chi^2$ P-value=0.02). However, no significant difference was observed for high CV ($Chi^2$ P-value > 0.05).

#### SM13. Comparisons of regional associations with previously identified pQTLs

We systematically evaluated whether the pQTL signals identified in our study had been previously reported in the literature. We extracted the main tables for three well-powered pQTL studies that used the SomaLogic platform in participants of European ancestries from deCODE[21], Fenland[22], and ARIC[23] studies. We also added a previous pQTL study conducted in the INTERVAL study using the 4K version of the SomaScan assay[24]. Finally, we considered the Sun et al UK Biobank study[25], which employed the Olink proteomic platform, and which represents the largest pQTL research study to date (54,219 individuals and 2,923 proteins). Although this study used a different platform, it provided an opportunity to compare and validate pQTL signals across different proteomic technologies.

To verify whether the pQTL signals identified in our study matched previously reported associations, we compared our regional associations against pQTLs from the above-mentioned studies. For each region associated with a specific protein, we used the boundaries of our loci to search for previously reported SNPs associated with that protein. In particular, for matches between our study and prior SomaLogic-based studies, if a SNP associated to the same aptamer or to a different aptamer targeting the same protein as our locus is located within its boundaries, it was marked as a match. For the UK Biobank study, which used the Olink platform, we compared regions based on UniProt annotations, ensuring comprehensive coverage of potential matches.

##### SM13.1 Reference pQTL studies and data harmonization

We are providing here the detailed methodology to build the reference file that we used to compare our results to those of 5 previous studies of high relevance for European pQTL studies.

deCODE[21]:

We reviewed the data provided in Supplementary Table 2 of the publication, which contains information on all 28,191 sentinel (primary) and secondary pQTL associations reported using genome build GRCh38. Since our analysis requires comparison with data based on genome build GRCh37, we performed liftover[26] to convert the coordinates accordingly.

Fenland[22]:

From the publication, we reviewed the data presented in Supplementary Table 2, which summarizes variant-protein target associations across 2,584 genomic regions. For our analysis, we considered all reported associations.

INTERVAL[24]:

We reviewed ST4-pQTL, which identified 1,927 primary significant associations (P-value<$1.5\times10^{-11}$) between 1,478 proteins and 764 genomic regions. Our analysis focused on the meta-analysis results.

ARIC[23]:

We extracted data from Supplementary Table 3.1, which reports aptamers with significant primary *cis*-pQTLs in individuals of European ancestry, and converted the genomic coordinates from genome build 38 to genome build GRCh37 using liftover[26].

UK Biobank[25]:

We extracted information from Supplementary Table 9, which lists primary significant associations (P-value<$1.7\times10^{-11}$) identified in the discovery cohort of European ancestry. The genomic coordinates were originally based on genome build GRCh38, and we performed liftover[26] to convert them to genome build GRCh37 for comparison with our data.

#### SM14. Identification of conditionally independent signals

To perform conditional analysis on the identified pQTLs, we used Genome-wide Complex Trait Analysis (GCTA) v.1.94.0.[27] First, each locus identified from the regional association step was extended by ±100 kb. Second, for each of these regions, we identified the set of conditionally independent SNPs by performing stepwise forward conditional regression using the *cojo-slct* function. We kept as conditionally independent SNPs all those with a joint P-value lower than *p1* ($5\times10^{-8}$/3,978) and an unconditional *P*-value lower than $1\times10^{-4}$. For each region, we generated a conditional dataset using the *cojo-cond* function, in which all the SNPs in the locus were conditioned on all identified independent SNPs. For the LD reference panel that is required by GCTA, we used the INTERVAL study data as the input.

#### SM15. Fine-mapping and colocalization of pQTLs

We performed fine-mapping of the conditionally independent signals to prioritise likely causal variants and then conducted colocalization across our pQTLs to identify signals that are potentially driven by shared causal variant(s). First, for each regional association and each conditionally independent SNP, we identified a credible set as the set of ranked variants with cumulative posterior inclusion probability (PIP) > 0.99 within the region. More precisely, to estimate the PIP for all variants in each conditional dataset, we initially estimated the phenotypic variance via the variance of each SNP's effect size from the unconditional model, the MAF, and the fixed sample size in GWAS meta-analysis via 'sdY.est' function in the coloc v5.2.3 R package. Then, using the estimated phenotypic variance as well as the SNP's conditional effect sizes and standard errors, we computed Approximate Bayes Factors (ABF) and calculated posterior probabilities by normalizing ABFs across variants. Variants were then ranked, and those with a cumulative posterior probability exceeding 0.99 were included in the 99% credible set.

Colocalization was tested only for pairs of conditionally independent signals whose 99% credible sets shared at least one variant. To meet the fundamental assumption of colocalization of only one causal variant per signal, we used the conditional datasets rather than the marginal summary, thus performing the colocalization tests for each pair of signals with overlapping credible-set variants. To implement colocalization analysis, we reformatted conditional datasets via the 'process.dataset' function and fed them into the 'coloc.abf' function, both available in coloc v5.2.3 R package with the default prior probability setting.[28] Colocalized pairs were identified when the posterior probability for hypothesis 4 (PP.H4) exceeded 0.80.

While individual examples can provide important biological insights, it is important to note that coloc's PP.H4 could be inflated for pQTL–pQTL colocalizations, as shared sample correlation can modestly bias results under the null. However, previous work[29] demonstrates that this inflation remains minor when trait–trait correlation is low, and simulation studies using HyPrColoc[30] confirm that ignoring sample overlap yields PP.H4 estimates closely matching fully adjusted models. Thus, strong colocalization signals—particularly when supported by independent biological evidence—can still be interpreted with confidence in this setting.

#### SM16. Modelling allelic complexity at pQTL loci

For each pQTL locus, the number of conditionally independent association signals was determined using stepwise conditional analysis as described above (**SM13)**. Loci were classified as primary if only one independent variant was detected and as secondary if two or more independent variants were identified. Logistic regression models were fitted with secondary signal status as the binary outcome. Predictor variables included protein panel status (newly added in SomaScan 7k vs. present in SomaScan 5k), locus category (*cis* or *trans*), and their interaction. Additional models were restricted to *cis* loci and adjusted for the absolute Z-score of the lead variant to account for differences in primary signal strength. To assess biological determinants of allelic complexity, further models included protein secretion pathway (intracellular, membrane, secreted) and log-transformed expected plasma protein concentration derived from the Human Protein Atlas. Models including protein concentration were fitted on complete-case subsets only, whereas models including protein location used the full dataset. All models were fitted using logistic regression, and results are reported as odds ratios with corresponding confidence intervals and Wald P-values.

#### SM17. Colocalization networks

To further investigate signal clustering within hotspots, we leveraged colocalization analysis results to pinpoint proteins potentially sharing regulatory processes within each hotspot. From each conditionally independent signal, we extracted the most significant conditional SNP from the corresponding conditional datasets. To ensure that colocalization between pairs was well represented by this SNP, we filtered out the colocalized pairs where LD between the most significant SNPs was below 0.8. he filtered colocalized pairs were then represented in a network framework, wherein each node denotes a variant-aptamer pair, and edges signify colocalization between nodes. Upon constructing these comprehensive networks for each hotspot, we applied community detection algorithms, a typical network analysis technique,[31] to extract maximal connected components of the network (clique), highlighting potential protein communities linked by shared causal variants, thus enabling detailed exploration of the biological pathways within each hotspot.

To prioritize biological signals to investigate, we performed a similar network clustering on robust (LD > 0.8 between index SNP) colocalizing signals outside hotspots.

#### SM18. pQTL–eQTL overlap and colocalization

##### SM18.1 Overlap of pQTLs and eQTLs

To check if the identified pQTLs are also eQTLs, we used available GTEx[32] v8 *cis*-eQTLs of European-ancestry participants in 49 tissues, lifted to GRCh37 using bcftools[26], lifting 99.6% of variants successfully. P-values and betas for all independent pQTL index variants were extracted from the eQTL data at exactly the chromosomal position for the respective alleles. We then checked the eQTLs for direction consistency and significance using a false discovery threshold (FDR) of ≤ 0.05.

##### SM18.2 Colocalization of pQTLs and eQTLs

To understand the extent to which genetic associations with protein levels might be regulated by altered gene expression, we tested all independent variants in the 7,870 *cis*- and *trans*-pQTL regions for colocalization with GTEx v8 *cis*-eQTLs for European-ancestry participants using the *coloc.abf()* function of the R-package coloc[28] using default priors and sdY=1. Colocalization was tested on the conditional summary statistics of the independent variants in the exact boundaries of the regional associations. Colocalizations with PPH4 ≥ 0.8 were considered significant. Summary statistics of available *cis*-eQTL regions for 49 tissues were obtained from GTEx[32] as described above. Sample size for the different tissues ranged from 130 for kidney cortex to 1176 for skeletal muscle (median=388, mean=522). In each region, for each set of conditioned pQTLs (conditioned on 12,095 independent index variants), pQTLs and eQTLs were matched by chromosome, genomic position, effect and non-effect allele. Then, each gene in the GTEx data with at least 50 matching variants[33] was tested for colocalization with the pQTL using Beta, SE, and P-value from the conditional analysis.
To assess tissue enrichment of colocalizations in *cis*-pQTLs, the expected number of significant colocalizations per tissue was computed as the total number of significant colocalizations over all tissues multiplied by the number of tested colocalizations in a tissue, divided by the total number of tested colocalizations. For each tissue, the enrichment ratio was computed as the number of significant colocalizations divided by the number of expected colocalizations, if the number of significant colocalizations per tissue was greater than the number of expected colocalizations for that tissue. The depletion ratio was calculated as minus the number of expected colocalizations divided by the number of true colocalizations, if the number of true colocalizations per tissue was smaller than the number of expected colocalizations. Thus, values > 1 indicate enrichment, and values < -1 indicate depletion. To compare colocalization patterns between proteins new in the 7k version and those previously assessed, we performed two Poisson regressions for *cis* and *trans*-pQTLs. For *cis*-p*QTLs*, we tested the difference in pQTLs that do and do not colocalize with the encoding gene in any tissue between the new and previously assessed proteins. For *trans*-pQTLs, we tested the

difference in the number of colocalizing genes per region between new and previously assessed proteins.

#### SM19. Mendelian Randomization association tests between plasma protein levels and phenotypes

##### SM19.1. Instruments selection

To test association between plasma protein levels and phenotypes through Mendelian Randomization (MR), we defined genetic instruments from the pQTL regional associations. To reduce the likelihood of horizontal pleiotropy in MR analyses, we used only the *cis* regional associations, thus considering for possible exposure only the aptamer-encoding gene pairs for which at least one *cis* significant signal was observed. For each *cis* signal, we selected the conditionally independent SNPs obtained through conditional analysis within each *cis* locus (extended by +/-100kb) associated with an aptamer, then extracted the corresponding effect sizes and standard errors for each SNP from the marginal associations. To avoid weak instruments, we excluded instruments with an F-statistic < 10, although none met this criterion. We further excluded 6 instrumental variables that were palindromic with intermediate allele frequencies. After filtering, 1,775 aptamers (1,778 aptamer–gene pairs) remained for analysis. The F-statistic was estimated as a function of PVE (proportion of variance explained[34]), sample size (N), and the number of instrumental variables (k), using the formula:

$$F = PVE\ (N - 1 - k)\ /\ (1 - PVE)k$$

where the PVE was computed as:

$$PVE\ =\ (2(Beta^2)MAF(1 - MAF))\ /(2(Beta^2)MAF(1 - MAF) + (Se^2)2(N)(MAF)(1 - MAF)))$$

##### SM19.2 Two-sample Mendelian Randomization analyses for diseases and traits

To identify the potential causal consequences of perturbing protein pathways, we performed Mendelian Randomization (MR), treating aptamers-encoding gene pairs as exposures and 2,003 harmonized diseases and traits as outcomes (**Supplementary Table 6**). Phenotype summary statistics were generated by the Million Veteran Program (MVP)[35] using fixed effects inverse-variance weighted GWAS meta-analysis of European-ancestry samples across MVP. The planned analytic sample consisted of approximately 22,000 participants, comprising overlapping groups with about 1,500 type 2 diabetes (T2D) cases, 1,500 cardiovascular disease (CVD) cases, 1,500 all-cause deaths, of which approximately 20% were expected to be attributable to coronary heart disease (CHD), 11,000 cancer cases, and 8,000-9,000 participants from a subcohort. Additional phenotype summary statistics were obtained from the UK Biobank (UKBB) and FinnGen[22], comprising more than 1.2 million participants in total. Phenotype harmonization and meta-analysis followed the MVP pipeline as referenced in Ferolito et al (2026)[36]. Briefly, disease endpoints were defined by mapping ICD-9/10 codes from VA electronic health records to Phecodes (≥2 ICD codes required for a case), with each *phe_XXX_YY* code (e.g., *phe_250_02*) analyzed as as a separate phenotype. Quantitative traits (laboratory measurements, vitals, anthropometrics) were curated separately and are not Phecode-based. For laboratory traits, MVP provided multiple phenotype representations (minimum, mean, maximum, and inverse-normal–transformed values), yielding up to six GWAS phenotypes per assay, each treated independently. Consistent with MVP, each phenotype representation was analyzed separately: for Phecodes, this means each distinct phe_XXX_YY entry; for laboratory and other non-Phecode traits, each unique phenotype (defined by lab × summary × transformation) or continuous trait was treated independently.

Cross-biobank harmonization across MVP, UKBB, and FinnGen used direct Phecode–Phecode matching, ICD-10→Phecode conversion in UKBB when necessary, and manual adjudication of phenotypes without direct matches. All harmonized phenotypes (both diseases and traits) were then assigned Experimental Factor Ontology (EFO) terms by MVP Ferolito et al[36]. For this manuscript, we used the harmonized phenotype labels and ontology assignments exactly as provided by MVP. We merged the Supplementary Table 1 from Ferolito et al[36] with per-biobank case and control counts and the MVP phenotype ontology table (provided by MVP with EFO ID, EFO term, and parent EFO term) into a unified dataset (**Supplementary Table 6**). To ensure complete merging of EFO terms, measurement-specific suffixes from laboratory traits (e.g., "_Max", "_Mean", "_INT") were removed, enabling a 100% match between our phenotype list and the MVP ontology file.

MR analyses were performed at the level of aptamer-gene-phenotype triplets. For triplets with a single genetic instrument (n = 1,512,297), we used the Wald ratio method. For triplets with multiple genetic instruments (n = 1,985,006), variants were combined using the inverse-variance weighted method implemented in TwoSampleMR.[37] Because variant availability differed across outcome GWAS datasets, the number of instruments available for a given aptamer–gene exposure could vary across phenotypes. If a chosen instrument was not directly available in a given outcome GWAS, a substituted proxy variant with $r^2 \geq 0.8$ (1000 Genomes EUR reference) was used, where available.

PHENOTYPE_CLASS is defined as "medications" if the phenotype name contains "Med", "diseases" if the phenotype has case–control structure (both cases and controls > 0 in any of MVP, UK Biobank, or FinnGen), and "traits" otherwise.

Signals that passed the MR multiple-testing threshold P-value < 0.05/(3,497,303) = $1.43 \times 10^{-8}$ were considered statistically significant. This Bonferroni-adjusted threshold accounts for the total number of unique aptamer–gene–phenotype triplets tested, which included 1,775 unique seqIDs, 1,543 unique UniProt IDs, and 2,003 unique phenotypes. All significant MR results are reported in **Supplementary Table 7**.

For MR-significant aptamer–gene–phenotype triplets, we additionally performed colocalization analyses between *cis*-pQTL summary statistics and the corresponding outcome GWAS signals using the coloc[28] package in R, following the general framework described in Ferolito et al. (2026).[36] Briefly, colocalization analyses were conducted using marginal (unadjusted) *cis*-pQTL summary statistics and GWAS summary statistics within a ±250-kb window around each MR instrument. Variants with minor allele frequency (MAF) > 1% were retained. For each tested region, posterior probabilities were estimated for the five colocalization hypotheses, and strong evidence of colocalization was defined as posterior probability for hypothesis 4 (PP.H4.abf), corresponding to a shared causal variant between the protein and phenotype association signals, greater than 0.8. Colocalization results, including maximum PP.H4.abf values across tested instruments, are reported in **Supplementary Table 7**. Pairs of significant gene-phenotype associations were systematically compared to previous findings from both eQTL and pQTL Mendelian Randomization of Ferolito et al (2026) (Supplementary table 10).[36]

##### SM19.3. Drug repurposing opportunities

We evaluated whether MR gene–trait associations recapitulate known drug indications, suggest repurposing opportunities, or identify potential indications for drugs under development. We used the gene–drug–indication table provided by the MVP consortium (derived from ChEMBL v34) and used in Ferolito et al. (2026)[36], which contains drugs, their molecular targets, and corresponding clinical indications and mode of actions. Each drug–indication pair is annotated with its maximum clinical development phase (max_phase_for_ind), and we retained drugs with max_phase_for_ind ≥ 3, corresponding to late-stage or approved compounds.

For each MR gene–trait pair, we searched for drugs targeting the corresponding gene and applied a hierarchical mapping to approved indications:
(i) exact EFO ID match,
(ii) exact EFO term match,
(iii) parent EFO match (EFO_Parent_ID),
(iv) phecode match (Genetic_phenotype_phecode), and
(v) expanded phenotype match (Matched_genetic_phenotype).

Associations with any (i–v) match were labeled “rediscovery”; if the gene had an approved drug but no matching Phase ≥ 3 indication, the pair was labeled a “repurposing candidate.” We recorded the tier producing the match in a match_type field. Genes without any drugs meeting the Phase ≥ 3 criterion were labeled “no drug–gene match.”

#### SM20. Functional consequence and epitope-binding annotations

Protein-altering variants may introduce aptamer-binding artifacts, leading to false associations between genetic variants and protein levels, which could bias causal estimates. To identify *cis*-pQTLs that could potentially be driven by aptamer-binding artifacts, we annotated conditionally independent variants (COJO SNPs) and their LD proxies ($r^2 > 0.8$, calculated in the INTERVAL dataset) using the Ensembl Variant Effect Predictor (VEP v113, GRCh37)[38]. Gene-level matching was performed using gene symbols to ensure alignment between VEP outputs and our protein annotations.

A pQTL was annotated as cis_epitope_effect = TRUE if all COJO variants (or at least one proxy per variant) had a ‘moderate’ or ‘high’ VEP consequence on the gene encoding the measured protein. This definition identifies pQTLs where there were no conditionally independent pQTLs that were free from potential epitope-binding artifacts. We included a second, more sensitive flag, cis_epitope_warning = TRUE, if any COJO variant (or proxy) in that pQTL had a ‘moderate’ or ‘high’ consequence on the gene encoding the measured protein.

In addition, we annotated general variant-level functional impact using VEP, also summarized at the locus level:

- impact_variant_vep = TRUE if any COJO variant (or proxy) had a moderate or high-impact consequence on any protein-coding gene.
- genes_with_consequence_types lists all Gene:ConsequenceType pairs (e.g., TP53:missense_variant) for moderate/high-impact annotations, filtered to protein-coding genes only.

All annotations were based on GRCh37 using the Ensembl VEP (custom GTF file from NCBI, GRCh37.p13), with a 500kb window. Default parameters were used unless otherwise specified.

#### SM21. LiftOver from GRCh37 into GRCh38

All analyses including the instrument selection are based on genome build GRCh37. The Mendelian Randomization and the colocalization analyses required to convert positions of the selected instruments to genome build GRCh38 to match MVP data for phenotype and GTEx data for eQTL. For this, we used the bcftools liftover tool[26].

We used liftover[26] to convert our results from the GRCh37 to the GRCh38 human genome assemblies. For our Mendelian randomization (MR) and colocalization analyses involving phenotypes, we utilized two human genome reference assemblies: the GRCh38 assembly obtained from the UCSC Genome Browser (http://hgdownload.soe.ucsc.edu/goldenPath/hg38/bigZips/hg38.fa.gz) and the GRCh37 assembly sourced from the 1000 Genomes Project repository (ftp://ftp.1000genomes.ebi.ac.uk/vol1/ftp/technical/reference/human_g1k_v37.fasta.gz).
To ensure accurate coordinate conversions between these two reference genomes, we employed chain files (hg19ToHg38.over.chain.gz). These chain files provide essential mappings of genomic regions, allowing us to translate variant coordinates effectively from the GRCh37 assembly to the GRCh38 assembly.

The first step in our analysis involved normalizing variant calls from the input VCF file, which was compressed in .gz format, using the GRCh37 reference genome. The normalization process included splitting multiallelic sites into biallelic records and compressing the output using bgzip. The command utilized for this normalization was:

```
bcftools norm -f {input.hg37} -c s -Oz -o {output.output_norm} {input.cojo_vcf}.gz
```

Following normalization, we performed a liftover of the normalized variant calls from the GRCh37 assembly to the GRCh38 assembly using the bcftools liftover command. This step involved the application of the following command:

```
bcftools +liftover --no-version -Ou {output.output_norm} -- -s {input.hg37} -f {input.hg38} -c {input.chain_file} > {output.output_vcf}
```

After successfully performing liftover, we converted the resultant VCF file into a plain text format for easier accessibility and analysis. This was achieved using the following command:

```
bcftools view {output.output_vcf} > {output.output_txt}
```

#### SM22. Per batch analysis

To examine whether the choice of combining batches in INTERVAL over meta-analysing them influenced our findings, we performed the following sensitivity analysis. We ran GWAS analyses independently in the two batches of the INTERVAL study, including the following covariates measured on 7,144 aptamers and 4,732 samples in batch 1 and 4,519 samples in batch 2: age, sex, time between blood draw and processing, season and the first 10 genetic principal components. Residuals were computed and used as input for Regenie as previously described. Meta-analysis was performed using METAL similarly to the main analysis but considering 3 studies: CHRIS, INTERVAL batch 1 and INTERVAL batch 2. Correlation of estimates was then systematically computed for the 7,144 aptamers. Correlation was very good, with a median of 0.94 and a minimum correlation of 0.82.
